# Mental disorders, receipt of acute cardiac care following myocardial infarction and the impact of the COVID-19 pandemic: a cohort study

**DOI:** 10.1101/2025.11.21.25340556

**Authors:** Kelly Fleetwood, John Nolan, Colin Berry, Debbie Cavers, Stewart W Mercer, Sandosh Padmanabhan, Daniel J Smith, Robert Stewart, Amanda Vettini, Caroline A Jackson, the CVD-COVID-UK/COVID-IMPACT Consortium

**Author notes:** Correspondence to: Kelly Fleetwood, Address: Usher Institute, Usher Building, The University of Edinburgh, 5‒7 Little France Road, Edinburgh BioQuarter ‒ Gate 3, Edinburgh, EH16 4UX, UK.

## Abstract

**Background and Aims:** People with a mental disorder have poorer myocardial infarction (MI) outcomes, with differences in cardiac care thought to be partly responsible. We compared receipt of guideline-informed acute MI care by mental disorder and assessed how the COVID-19 pandemic affected associations.

**Methods:** We identified people with MI in England (November 2019 - February 2023) from the Myocardial Ischaemia National Audit Project (MINAP), ascertaining prior mental disorder from linked hospitalisation and primary care records and extracting care standards from MINAP. We used logistic regression to compare care standards for ST-elevation MI (STEMI) and non-STEMI (NSTEMI) between people with each of schizophrenia, bipolar disorder or depression versus those without any of these disorders, adjusting for confounders and investigating differences over time.

**Results:** We included 131,075 NSTEMI and 79,045 STEMI cases. For NSTEMI, people with prior mental disorder had lower odds of angiography eligibility and receipt, cardiac ward admission and cardiac rehabilitation referral. Odds ratios (95% CIs) ranged from 0.25 (0.20, 0.31) for angiography receipt for schizophrenia to 0.92 (0.89, 0.96) for cardiac ward admission for depression. For STEMI, there was no evidence of care differences for depression; however, people with bipolar disorder were less likely to meet call-to-balloon targets and people with schizophrenia were less likely to be referred for cardiac rehabilitation and receive indicated secondary prevention medication. Disparities were generally unaffected by the COVID-19 pandemic.

**Conclusions:** People with a mental disorder are less likely to receive guideline-informed MI care, with variation by MI type, care standard and mental disorder.

**Summary:** We used linked electronic health records from over 200 000 NSTEMI and STEMI patients in England to compare receipt of guideline-informed care for myocardial infarction by mental disorder status and assess how the COVID-19 pandemic affected associations. We identified disparities in a range of care standards for both NSTEMI and STEMI, which were greatest for people with schizophrenia, but also evident for bipolar disorder and depression. Disparities were generally unaffected by the COVID-19 pandemic.

## Introduction

Cardiovascular disease is a major contributor to the entrenched disparity in life expectancy of people with a mental disorder compared to the general population.^1–6^ Mental disorders, such as schizophrenia, bipolar disorder and depression, are associated with a higher incidence of major cardiovascular events, including myocardial infarction (MI).^7–9^ Following MI, pre-existing mental disorder is associated with higher 30-day and 1-year mortality and greater risk of further vascular events.^10,11^ There is growing evidence of mental health disparities in receipt of clinical care for physical conditions, including cardiovascular disease, which may partly account for differences in outcomes. A recent systematic review reported lower rates of angiography, percutaneous coronary intervention (PCI) and receipt of secondary prevention medication following MI in people with, compared to without, a clinically diagnosed mental disorder.^10^ However, existing studies are generally non-contemporary, have rarely reported findings separately for ST-elevation MI (STEMI) and non-ST-elevation MI (NSTEMI) and have largely focused on receipt of procedures. To our knowledge, no study has comprehensively compared receipt of guideline-informed clinical care for acute management of MI by mental disorder. Important elements of clinical care include early management, treatment and early initiation of rehabilitation, to improve chances of recovery and survival following MI. Examination of such care standards by mental disorder could provide detailed insight into inequalities across the acute clinical care pathway.

Delivery of health care, including cardiac care, was disrupted in the UK during the COVID-19 pandemic.^12^ Reports on whether this impacted MI care standard targets in the general population are conflicting.^13,14^ No study, to our knowledge, has examined the impact of the pandemic on mental health disparities in acute cardiac care. It is important to examine whether marginalised groups experiencing MI were disproportionately affected by the COVID-19 pandemic in order to address existing inequalities and better inform responses to future disruptions to delivery of care.

This study aimed to determine (1) how acute care for MI in England differs by mental disorder status, by comparing receipt of clinical care standards for NSTEMI and STEMI among those with versus without schizophrenia, bipolar disorder or depression, using contemporary data, and (2) the impact of COVID-19 on the associations between mental disorder and receipt of acute care for MI.

## Methods

We have reported this study in accordance with the Reporting of Studies Conducted using Observational Routinely-Collected Health Data (RECORD) statement.^15^

### Setting and data sources

We accessed national linked electronic health records from England in NHS England’s (NHSE) Secure Data Environment (SDE), made available through the British Heart Foundation Data Science Centre’s <u>CVD-COVID-UK/COVID-IMPACT Consortium</u>.^16^ Our primary data source was the Myocardial Ischaemia National Audit Project (MINAP), a clinical audit of patients admitted to hospital with MI, which includes information on care from the call for help to discharge, patient demographics, medical history and clinical characteristics of the MI.^17^ We also used the General Practice Extraction Service (GPES) Data for Pandemic Planning and Research (GDPPR) dataset, which captures data from 98% of GP practices in England (Supplementary data online, *Methods S1*).^18^ The GDPPR dataset includes patients who are currently registered and deceased patients with a date of death on or after 1 November 2019, with records at time of data extraction available up to 31 July 2024. GP records transfer between practices when a patient moves and so should capture an individual’s entire medical history. We also used the Hospital Episode Statistics Admitted Patient Care (HES APC) dataset, which included all inpatient admissions and day cases from 1 April 1997 to 31 July 2024. We also used data from: COVID-19 Hospitalisations in England Surveillance System, Second Generation Surveillance System and Secondary Uses Service, all of which can be used to identify people with COVID-19 infection.

### Study design and population

We conducted a retrospective cohort study, including all adults (≥ 18 years of age) with a final diagnosis of NSTEMI or STEMI recorded in MINAP from 1 November 2019 to 28 February 2023. We did not include events prior to 1 November 2019, since GP data was unavailable for people who died before this date. We excluded patients without a linked GDPPR record and those who failed quality assurance checks. For each individual we included the first record of an MI within the study period.

### Mental disorder

We ascertained history of schizophrenia, bipolar disorder and unipolar depression (of any severity) from the GDPPR and HES APC datasets, based on diagnoses at any time prior to each individual’s date of MI admission. We defined mutually exclusive groups of people with schizophrenia, bipolar disorder or depression using a severity hierarchy, with schizophrenia considered the most severe, followed by bipolar disorder and depression. For the HES APC dataset, we identified each disorder from the primary and secondary diagnosis fields, using ICD-10 codes for schizophrenia (F20, F25), bipolar disorder (F30, F31) and depression (F32, F33). For the GDPPR dataset, we identified each disorder using SNOMED codes (Supplementary data online, *Methods S1*). The comparison group included people without a GDPPR or HES APC record of schizophrenia, bipolar disorder or depression prior to their date of MI admission.

### Standards of care for NSTEMI and STEMI

Our outcomes were based on the quality improvement metrics that are measured in MINAP annual reports,^19^ which are based on national and international clinical guidelines.^20,21^ We assessed the following care standards relevant for NSTEMI: eligibility for angiography (as defined by the clinical team; reasons for ineligibility could include advanced malignancy, dementia, progressive neurological disease, or other clinical reasons); receipt of angiography (amongst people eligible for angiography); angiography within 72 hours (amongst people who received angiography); and admission to a cardiac ward) defined as coronary care unit or cardiac ward (amongst people who did not die in accident and emergency). We examined the following care standards for STEMI amongst people who received primary PCI: call-to-balloon (CTB) time (i.e. time from call for help to receipt of primary PCI) within 120 minutes; CTB time within 150 minutes; door-to-balloon (DTB) time (i.e. time from arrival at hospital to receipt of primary PCI) within 60 minutes; and DTB time within 90 minutes. We also assessed two care outcomes relevant to both STEMI and NSTEMI amongst people discharged home: indicated secondary prevention medication prescribed at discharge; and referral for cardiac rehabilitation (Supplementary data online, *Methods S2*). Whilst we did not analyse receipt of reperfusion in statistical models, we summarised the proportion receiving PCI for NSTEMI and primary PCI for STEMI by mental disorder status, for completeness.

### Covariates

We ascertained age, sex, deprivation (based on quintiles of the Index of Multiple Deprivation,^22^ a neighbourhood-level measure of deprivation) and ethnicity from the CVD-COVID-UK/COVID-IMPACT Consortium key patient characteristics table.^23^ We defined MI admission timing variables: daytime or overnight; weekday or weekend and 4-month period during which the admission occurred; and identified the hospital where the patient was admitted (Supplementary data online, *Methods S3*).

We identified history of MI and comorbid angina, cerebrovascular disease, chronic renal failure, diabetes and heart failure from the MINAP dataset and from diagnoses recorded in GDPPR or HES APC prior to the date of MI admission (Supplementary data online, *Methods S3*)^16,24–28^. We identified history of PCI from the MINAP dataset and from procedures recorded in HES APC^11^ (Supplementary data online, *Methods S3*). We ascertained MI presentation features including cardiac arrest, cardiogenic shock, ST deviation, creatinine within the first 24 hours after admission (μmol/L), heart rate on admission and the first systolic blood pressure (SBP) (mmHg) measurement after admission from the MINAP dataset. We identified patients who had COVID-19 within 14 days either side of their MI (Supplementary data online, *Methods S3*).

### Statistical analyses

We analysed NSTEMI and STEMI separately, using mixed-effects logistic regression to obtain odds ratios for the receipt of each standard of care among patients with each of schizophrenia, bipolar disorder or depression versus patients without any of these disorders. For each of NSTEMI and STEMI, we serially adjusted for groups of potential confounding factors: model 1 adjusted for age and sex, with random effects for the intercept for each hospital; model 2 additionally adjusted for timing variables; model 3 additionally adjusted for ethnicity and deprivation; model 4 additionally adjusted for clinical history (history of MI, history of PCI and comorbidities); and model 5 additionally adjusted for MI presentation features (cardiac arrest, cardiogenic shock, creatinine, heart rate, SBP and COVID-19 status for both types of MI, and additionally ST deviation for NSTEMI only). Age, creatinine, heart rate and SBP were included in the models as continuous variables. We scaled age, SBP and heart rate by subtracting their respective means and dividing by their standard deviations. Creatinine was right skewed and hence was log transformed prior to scaling to improve convergence of the models. For all continuous variables, we included both linear and quadratic terms in the models because exploratory plots suggested that quadratic functions were sufficient for describing the relationships between these variables and the outcomes. We investigated whether any associations between mental disorders and receipt of standards of care were affected by the response to the COVID-19 pandemic. Given that our study data was only available from November 2019 we evaluated the effect of the pandemic by comparing the associations in the pre-pandemic period (November 2019 – February 2020) to the associations in each of the following 4-month periods. We categorised time into four-month intervals because we wanted to capture fluctuations in care throughout our study period without making prior assumptions that specific periods (for example lockdown periods) would have better or worse attainment of care standards. In addition to lockdowns, receipt of care throughout our study period may have been affected by periods of increased or decreased demand on ambulance services and hospitals. To compare the associations in the pre-pandemic period to those in later periods, we refitted the model using a binary mental disorder variable (any of schizophrenia, bipolar disorder or depression versus none of these disorders) since there were insufficient numbers to investigate the interaction for individual mental disorders. We then fitted a further model additionally including the interaction between the binary mental disorder variable and 4-month time period. We used a likelihood ratio test to evaluate whether the model with the interaction provided a better fit to the data than the model without the interaction.

The primary analyses excluded patients with missing data in the outcome or any of the covariates. Missing data was minimal amongst the sociodemographic, timing and clinical history covariates, but higher amongst the MI presentation covariates. Hence, we conducted a post hoc sensitivity analysis based on a larger set of patients, excluding only those with missing data in the outcome or the sociodemographic, timing and clinical history covariates. This analysis included models 1 to 4 as above, and tested the interaction based on model 4 (again using the binary mental disorder variable for evaluating the interaction).

In accordance with statistical disclosure control rules, counts are rounded to the nearest 5 and counts less than 10 are suppressed. We used R (version 4.1.3 and above)^29^ to conduct the statistical analysis and specifically the R package lme4 (version 1.1.35.3 and above)^30^ to fit mixed-effects logistic regression models. All R code is available in our GitHub repository (https://github.com/BHFDSC/CCU046_01).

## Results

### Descriptive findings

Our study population included 210,120 people with a final diagnosis of MI recorded in MINAP between 1 November 2019 and 28 February 2023 (Figure 1, Supplementary data online, *Figure S1*).

**Figure 1:**
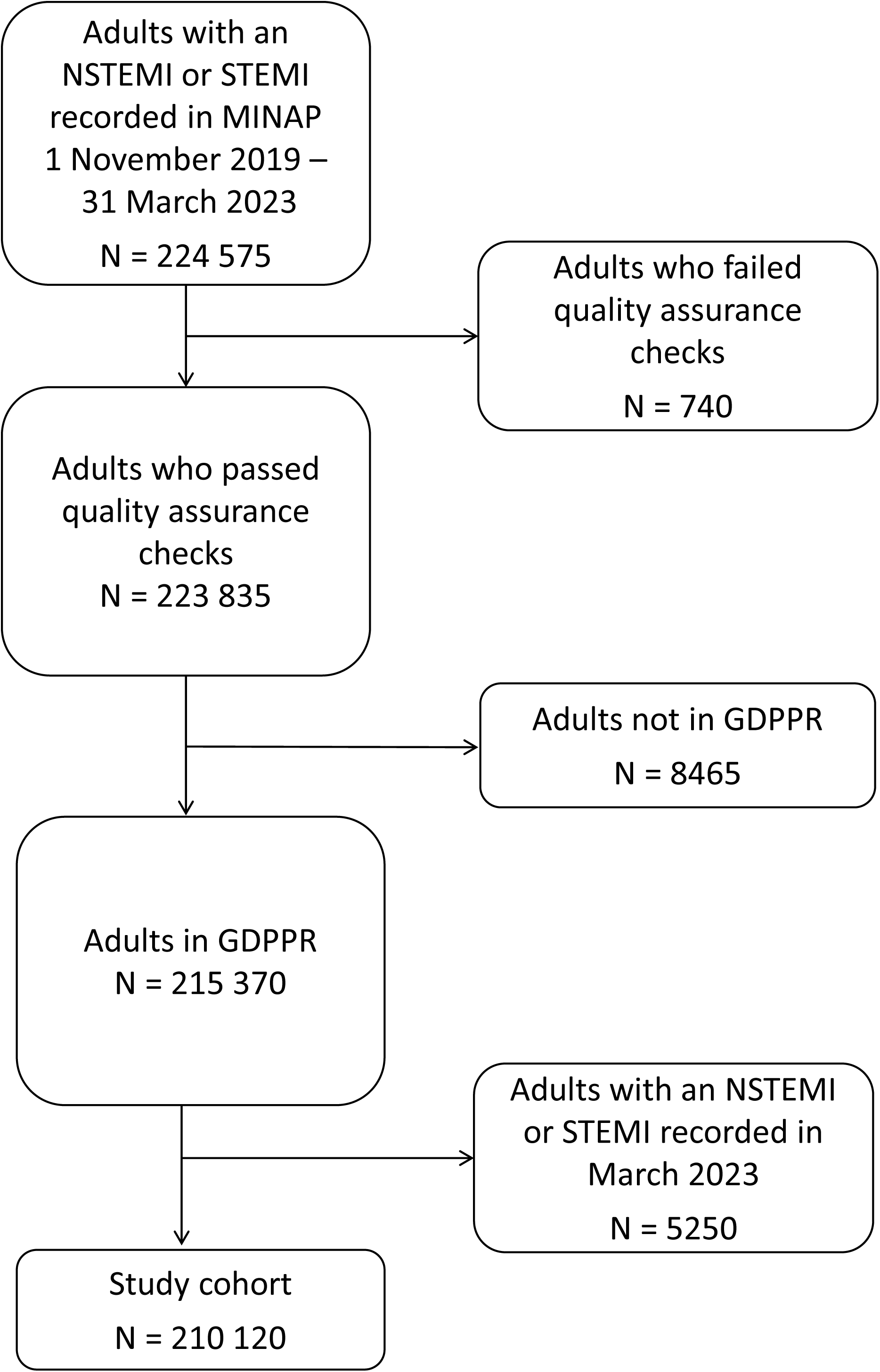
Flow diagram for the study cohort

Among 131,075 people with an NSTEMI, 875 (0.7%) had schizophrenia, 750 (0.6%) had bipolar disorder and 34,610 (26.4%) had depression. Among 79,045 people with a STEMI event, 490 (0.6%) had schizophrenia, 405 (0.5%) had bipolar disorder and 19,230 (24.3%) had depression. Relative to the comparison group, people with each mental disorder were younger at the time of MI, more likely to be female and more likely to live in deprived areas. The proportion of patients in ethnic minority groups was over-represented among those with schizophrenia. In general, for both NSTEMI and STEMI, the proportion of people with prior comorbidities was higher in those with mental disorder, but MI presentation features, including COVID-19 infection status, generally did not differ by mental disorder status (Tables 1 and 2).

**Table 1.** Baseline characteristics of patients with a MINAP record of an NSTEMI, by mental disorder status^a^.

| Characteristic | Schizophrenia<br>(N = 875) | Bipolar disorder<br>(N = 750) | Depression<br>(N = 34 610) | None of these disorders<br>(N =94 840) |
| --- | --- | --- | --- | --- |
| **Sociodemographics** |  |  |  |  |
| Age at MI (years), mean (SD) | 64.7 (12.5) | 65.3 (12.9) | 67.5 (13.0) | 70.6 (13.3) |
| Female (%) | 295 (33.7) | 320 (42.7) | 15 170 (43.8) | 27 840 (29.4) |
| IMD decile (%) |  |  |  |  |
| 1 Most deprived | 195 (22.3) | 120 (16.0) | 5210 (15.1) | 9710 (10.2) |
| 2 | 155 (17.7) | 105 (14.0) | 4155 (12.0) | 9150 (9.6) |
| 3 | 115 (13.1) | 75 (10.0) | 3830 (11.1) | 9290 (9.8) |
| 4 | 105 (12.0) | 105 (14.0) | 3655 (10.6) | 9570 (10.1) |
| 5 | 70 (8.0) | 65 (8.7) | 3470 (10.0) | 9620 (10.1) |
| 6 | 65 (7.4) | 70 (9.3) | 3275 (9.5) | 9805 (10.3) |
| 7 | 45 (5.1) | 75 (10.0) | 3100 (9.0) | 9535 (10.1) |
| 8 | 55 (6.3) | 45 (6.0) | 2870 (8.3) | 9425 (9.9) |
| 9 | 35 (4.0) | 55 (7.3) | 2645 (7.6) | 9210 (9.7) |
| 10 least deprived | 30 (3.4) | 40 (5.3) | 2380 (6.9) | 9410 (9.9) |
| Missing | <10 (<1.1) | <10 (<1.3) | 25 (0.1) | 110 (0.1) |
| Ethnicity |  |  |  |  |
| Black | 30 (3.4) | 10 (1.3) | 365 (1.1) | 1700 (1.8) |
| South Asian | 125 (14.3) | 50 (6.7) | 1785 (5.2) | 7990 (8.4) |
| White | 680 (77.7) | 675 (90.0) | 31 535 (91.1) | 81 330 (85.8) |
| Mixed | <10 (<1.1) | <10 (<1.3) | 240 (0.7) | 685 (0.7) |
| Other ethnic groups | 30 (3.4) | 10 (1.3) | 675 (2.0) | 2970 (3.1) |
| Missing | <10 (<1.1) | <10 (<1.3) | 10 (<0.1) | 170 (0.2) |
| **Location and timing** |  |  |  |  |
| NHS trust |  |  |  |  |
| Available | 855 (97.7) | 725 (96.7) | 33 660 (97.3) | 92 260 (97.3) |
| Missing | 20 (2.3) | 25 (3.3) | 955 (2.8) | 2580 (2.7) |
| Admission day |  |  |  |  |
| Week day | 670 (76.6) | 565 (75.3) | 26 030 (75.2) | 71 370 (75.3) |
| Weekend | 205 (23.4) | 190 (25.3) | 8580 (24.8) | 23 470 (24.7) |
| Admission timing |  |  |  |  |
| Daytime | 485 (55.4) | 425 (56.7) | 20 815 (60.1) | 58 720 (61.9) |
| Overnight | 390 (44.6) | 330 (44.0) | 13 795 (39.9) | 36 120 (38.1) |
| **Clinical history** |  |  |  |  |
| Previous MI (%) | 235 (26.9) | 215 (28.7) | 9220 (26.6) | 21 755 (22.9) |
| Previous PCI (%) | 200 (22.9) | 210 (28.0) | 10 525 (30.4) | 27 310 (28.8) |
| Angina (%) | 420 (48.0) | 390 (52.0) | 17 200 (49.7) | 42 710 (45.0) |
| Cerebrovascular disease (%) | 225 (25.7) | 195 (26.0) | 7105 (20.5) | 14 980 (15.8) |
| Chronic renal failure (%) | 100 (11.4) | 85 (11.3) | 3455 (10.0) | 9750 (10.3) |
| Diabetes (%) | 430 (49.1) | 320 (42.7) | 13 780 (39.8) | 33 475 (35.3) |
| Heart failure (%) | 300 (34.3) | 215 (28.7) | 10 030 (29.0) | 25 700 (27.1) |
| **MI presentation features** |  |  |  |  |
| ST deviation (%) | 170 (19.4) | 130 (17.3) | 6585 (19.0) | 19 320 (20.4) |
| Missing (%) | 30 (3.4) | 15 (2.0) | 1140 (3.3) | 2810 (3.0) |
| Cardiogenic shock (%) | <10 (<1.1) | <10 (<1.3) | 125 (0.4) | 410 (0.4) |
| Missing (%) | 115 (13.1) | 75 (10.0) | 3820 (11.0) | 10 310 (10.9) |
| Cardiac arrest (%) | 35 (4.0) | 15 (2.0) | 755 (2.2) | 2445 (2.6) |
| Missing (%) | 25 (2.9) | 10 (1.3) | 660 (1.9) | 1880 (2.0) |
| Creatinine (μmol/L) |  |  |  |  |
| Median [IQR] | 83.0 [68.0, 104.0] | 83.0 [68.0, 104.0] | 82.0 [68.0, 101.0] | 85.0 [72.0, 105.0] |
| Missing (%) | 35 (4.0) | 20 (2.7) | 970 (2.8) | 2475 (2.6) |
| SBP (mmHg) |  |  |  |  |
| Mean (SD) | 138.2 (27.5) | 140.0 (28.2) | 143.3 (27.3) | 145.1 (27.4) |
| Missing (%) | 35 (4.0) | 15 (2.0) | 795 (2.3) | 2340 (2.5) |
| Heart rate (bpm) |  |  |  |  |
| Mean (SD) | 85.8 (20.4) | 82.8 (19.8) | 80.7 (19.0) | 79.8 (19.2) |
| Missing (%) | 30 (3.4) | 15 (2.0) | 830 (2.4) | 2420 (2.6) |
| COVID-19 positive (%) | 50 (5.7) | 45 (6.0) | 1715 (5.0) | 4995 (5.3) |

<sup>a</sup>In accordance with NHSE's SDE statistical disclosure control rules, counts are rounded to the nearest 5 and counts less than 10 are suppressed

**Table 2.**
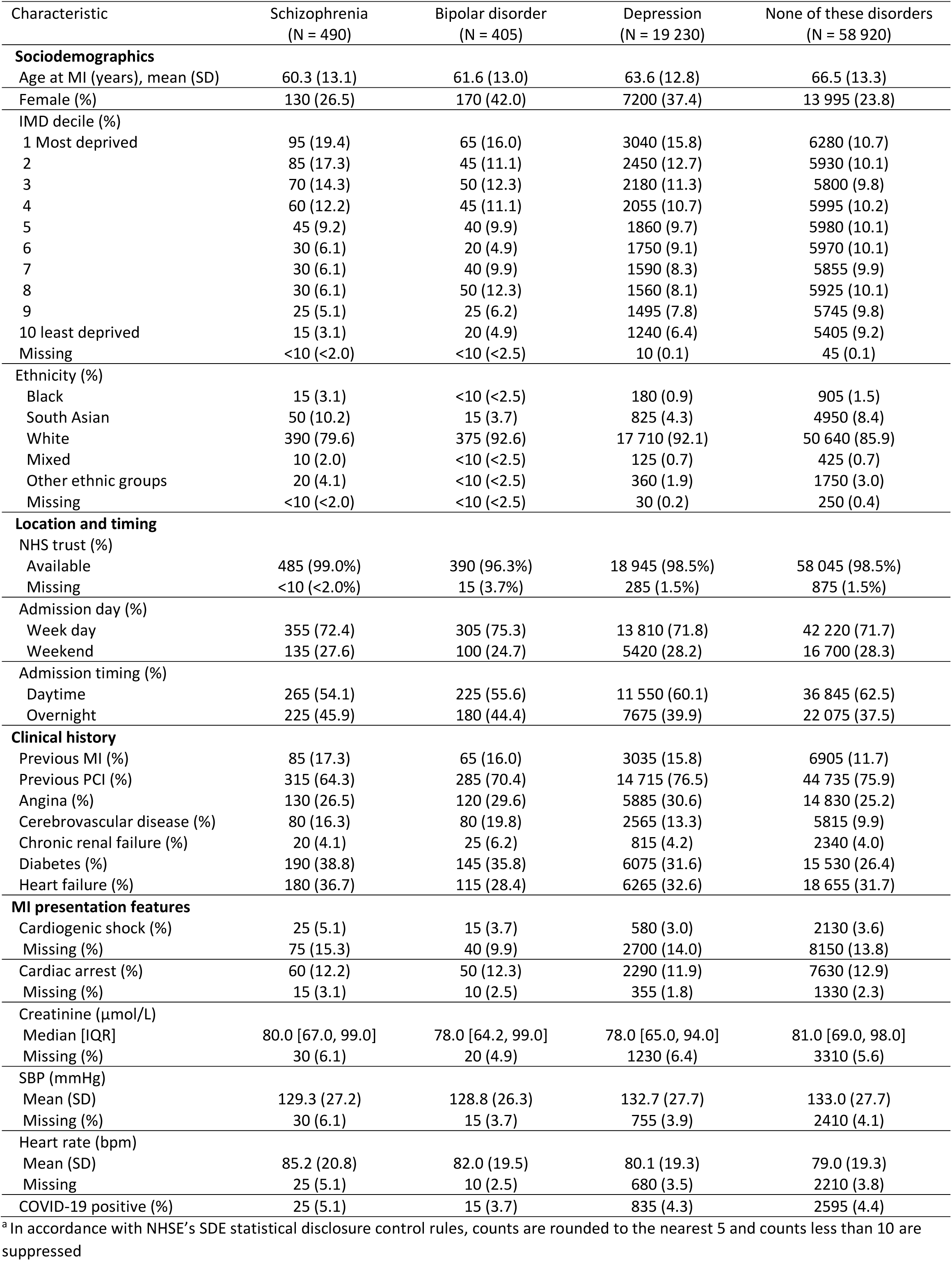
Baseline characteristics of patients with a MINAP record of a STEMI, by mental disorder status^a^.

Relative to the comparison group, the proportion of patients receiving each NSTEMI care standard was lower in those with a mental disorder, with the lowest proportions in people with schizophrenia (Table 3). Patterns of receipt of care by mental disorder for STEMI were more mixed, with variation by individual disorder and across care standards (Table 3).

**Table 3.** Receipt of care among people with each of NSTEMI and STEMI, by mental disorder status^a^.

| Receipt of clinical care | Schizophrenia | Bipolar disorder | Depression | None of these disorders |
| --- | --- | --- | --- | --- |
| **NSTEMI** |  |  |  |  |
| Angiography eligibility (%) |  |  |  |  |
| N (all patients) | 875 | 750 | 34 610 | 94 840 |
| Yes | 690 (78.9) | 610 (81.3) | 28 605 (82.6) | 79 290 (83.6) |
| No | 145 (16.6) | 100 (13.3) | 4310 (12.5) | 11 400 (12.0) |
| Missing | 45 (5.1) | 40 (5.3) | 1695 (4.9) | 4150 (4.4) |
| Angiography receipt (%) |  |  |  |  |
| N (patients eligible for angiography) | 690 | 610 | 28 605 | 79 290 |
| Yes | 490 (71.0) | 510 (83.6) | 24 580 (85.9) | 68 575 (86.5) |
| No | 140 (20.3) | 80 (13.1) | 3365 (11.8) | 9325 (11.8) |
| Patient refused | 60 (8.7) | 20 (3.3) | 660 (2.3) | 1390 (1.8) |
| Angiography within 72 hours (%) |  |  |  |  |
| N (patients who received angiography) | 490 | 510 | 24 580 | 68 575 |
| Yes | 220 (44.9) | 235 (46.1) | 11 765 (47.9) | 33 835 (49.3) |
| No | 175 (35.7) | 175 (34.3) | 8230 (33.5) | 22 885 (33.4) |
| Missing | 95 (19.4) | 100 (19.6) | 4585 (18.7) | 11860 (17.3) |
| Admission to cardiac ward (%) |  |  |  |  |
| N (patients who did not die in A&E) | 875 | 750 | 34 595 | 94 750 |
| Yes | 335 (38.3) | 285 (38.0) | 14 595 (42.2) | 40 840 (43.1) |
| No | 510 (58.3) | 440 (58.7) | 19 150 (55.4) | 51 555 (54.4) |
| Missing | 25 (2.9) | 25 (3.3) | 845 (2.4) | 2355 (2.5) |
| Referred for cardiac rehabilitation (%) |  |  |  |  |
| N (patients discharged home and without referral for cardiac rehabilitation recorded as 'Not indicated'<sup>b</sup>) | 580 | 480 | 24 300 | 66 345 |
| Yes | 420 (72.4) | 385 (80.2) | 20 265 (83.4) | 55 435 (83.6) |
| No | 80 (13.8) | 45 (9.4) | 2125 (8.7) | 5845 (8.8) |
| Patient declined | 20 (3.4) | 10 (2.1) | 380 (1.6) | 820 (1.2) |
| Missing | 55 (9.5) | 40 (8.3) | 1535 (6.3) | 4250 (6.4) |
| Receipt of indicated secondary prevention medication (%) |  |  |  |  |
| N (patients discharged home) | 660 | 535 | 26 210 | 71 420 |
| Yes | 535 (81.1) | 440 (82.2) | 21 830 (83.3) | 60 200 (84.3) |
| No | 100 (15.2) | 80 (15.0) | 3630 (13.8) | 9410 (13.2) |
| Missing | 25 (3.8) | 15 (2.8) | 750 (2.9) | 1810 (2.5) |
| **STEMI** |  |  |  |  |
| Call to balloon within 120 minutes (%) |  |  |  |  |
| N (patients who underwent primary PCI) | 355 | 315 | 15 655 | 48 015 |
| Yes | 110 (31.0) | 65 (20.6) | 4475 (28.6) | 14 270 (29.7) |
| No | 190 (53.5) | 210 (66.7) | 9395 (60.0) | 27 980 (58.3) |
| Missing | 55 (15.5) | 40 (12.7) | 1785 (11.4) | 5770 (12.0) |
| Call to balloon within 150 minutes (%) |  |  |  |  |
| N (patients who underwent primary PCI) | 355 | 315 | 15 655 | 48 015 |
| Yes | 175 (49.3) | 140 (44.4) | 8175 (52.2) | 25 280 (52.7) |
| No | 125 (35.2) | 135 (42.9) | 5695 (36.4) | 16 965 (35.3) |
| Missing | 55 (15.5) | 40 (12.7) | 1785 (11.4) | 5770 (12.0) |
| Door to balloon within 60 mins (%) |  |  |  |  |
| N (patients who underwent primary PCI) | 355 | 315 | 15 655 | 48 015 |
| Yes | 55 (15.5) | 35 (11.1) | 2005 (12.8) | 6340 (13.2) |
| No | 235 (66.2) | 230 (73.0) | 11 385 (72.7) | 34 080 (71.0) |
| Missing | 65 (18.3) | 50 (15.9) | 2260 (14.4) | 7595 (15.8) |
| Door to balloon within 90 mins (%) |  |  |  |  |
| N (patients who underwent primary PCI) | 355 | 315 | 15 655 | 48 015 |
| Yes | 155 (43.7) | 130 (41.3) | 6800 (43.4) | 20 830 (43.4) |
| No | 135 (38.0) | 140 (44.4) | 6595 (42.1) | 19 590 (40.8) |
| Missing | 65 (18.3) | 50 (15.9) | 2260 (14.4) | 7595 (15.8) |
| Referral for cardiac rehabilitation (%) |  |  |  |  |
| N (patients discharged home and without referral for cardiac rehabilitation recorded as 'Not indicated') | 360 | 320 | 15 965 | 48 720 |
| Yes | 320 (88.9) | 290 (90.6) | 14 805 (92.7) | 44 980 (92.3) |
| No | 25 (6.9) | 15 (4.7) | 490 (3.1) | 1730 (3.6) |
| Patient declined | 10 (2.8) | <10 (<3.1) | 100 (0.6) | 265 (0.5) |
| Missing | 10 (2.8) | 15 (4.7) | 575 (3.6) | 1745 (3.6) |
| Receipt of indicated secondary prevention medication (%) |  |  |  |  |
| N (patients discharged home) | 380 | 330 | 16 300 | 49 795 |
| Yes | 340 (89.5) | 305 (92.4) | 15 085 (92.5) | 46 205 (92.8) |
| No | 25 (6.6) | 15 (4.5) | 805 (4.9) | 2525 (5.1) |
| Missing | 15 (3.9) | <10 (<3.0) | 410 (2.5) | 1065 (2.1) |

<sup>a</sup>In accordance with NHSE's SDE statistical disclosure control rules, counts are rounded to the nearest 5 and counts less than 10 are suppressed
<sup>b</sup>Out of NSTEMI patients discharged home, referral for cardiac rehabilitation was recorded as 'Not indicated' for 12.9% of patients with schizophrenia, 9.3% of patients with bipolar disorder, 7.3% of patients with depression and 7.1% of patients without any of these disorders.
<sup>c</sup>Out of STEMI patients discharged home, referral for cardiac rehabilitation was recorded as 'Not indicated' for 5.3% of patients with schizophrenia, <3.0% of patients with bipolar disorder, 2.1% of patients with depression and 2.2% of patients without any of these disorders.

### Mental disorder and receipt of care for NSTEMI

Logistic regression indicated that people with a mental disorder were less likely to receive most NSTEMI care standards, with differences largest for those with schizophrenia (Figure 2). For each mental disorder, the effect estimates from models one to three were similar (Supplementary data online, *Figure S2)*, with additional adjustment for clinical history and MI presentation leading to some attenuation. Based on the fully adjusted models, mental disorder was associated with lower odds of being considered eligible for angiography (schizophrenia: OR 0.41, 95% CI 0.32, 0.54; bipolar disorder: OR 0.74, 95% CI 0.54, 1.00; depression: OR 0.78, 95% CI 0.74, 0.83) and, amongst those eligible, to then receive angiography (schizophrenia: OR 0.25, 95% CI 0.20, 0.31; bipolar disorder: OR 0.58, 95% CI 0.44, 0.76; depression: OR 0.81, 95% CI 0.77, 0.85). There was no clear difference in receipt of angiography within 72 hours by mental disorder status (Figure 2).

**Figure 2:**
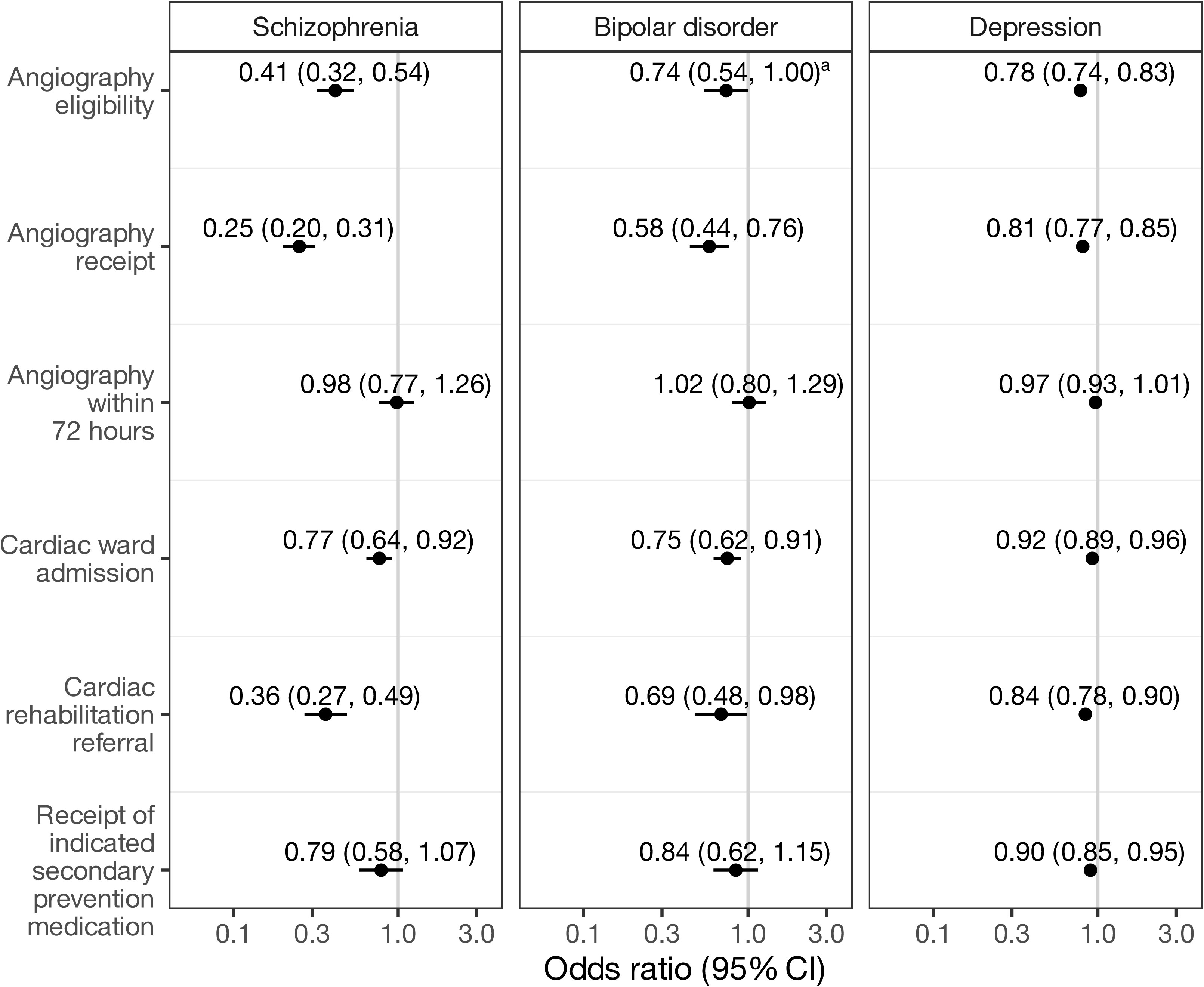
Odds ratios for receipt of each care standard following NSTEMI in patients with schizophrenia, bipolar disorder or depression versus those without any of these disorders. Odds ratios are adjusted for sociodemographic characteristics, hospital, MI timing, comorbidities and MI presentation features (model 5).

Among patients who did receive angiography, the proportion receiving PCI was lower in those with mental disorder, particularly schizophrenia (45.9%), versus the comparison group (55.9%), whilst the proportion for whom PCI was recorded as not performed or arranged was higher in those with a mental disorder (Supplementary data online, *Table S1*).

Logistic regression indicated that people with a mental disorder were less likely to be admitted to a cardiac ward and to be referred for cardiac rehabilitation. Odds of receiving indicated secondary prevention medication were lower in people with depression than the comparison group but there was not a statistically significant difference for people with schizophrenia or bipolar disorder (Figure 2).

### Mental disorder and receipt of care for STEMI

Around 75% of people with schizophrenia or bipolar disorder received initial reperfusion treatment relative to around 80% amongst those with depression or in the comparison group (Supplementary data online, *Table S2*).

Based on logistic regression models, associations between mental disorder status and receipt of guideline-informed care standards for STEMI differed by individual mental disorder and care indicator. In general, additional adjustment for covariates only minimally altered effect estimates (Supplementary data online, *Figure S3)*. Among patients who received a PCI, based on the fully adjusted models, bipolar disorder was associated with 42% (OR 0.58, 95% CI 0.42, 0.79) lower odds of meeting the 120-minute CTB target and 28% (OR 0.72, 95% CI 0.55, 0.93) lower odds of meeting the 150-minute CTB target. There was no association between either schizophrenia or depression and CTB time (Figure 3). We also observed no statistically significant difference in the achievement of the 60-minute or 90-minute DTB target between patients with versus without each mental disorder. Relative to the comparison group, patients with schizophrenia were less likely to be referred for cardiac rehabilitation (OR 0.38, 95% CI 0.23, 0.61) and to receive indicated secondary prevention medication (OR 0.46, 95% CI 0.27, 0.77; Figure 3). However, there was no difference in referral to cardiac rehabilitation or receipt of secondary prevention medication among those with each of bipolar disorder and depression versus the comparison group.

**Figure 3:**
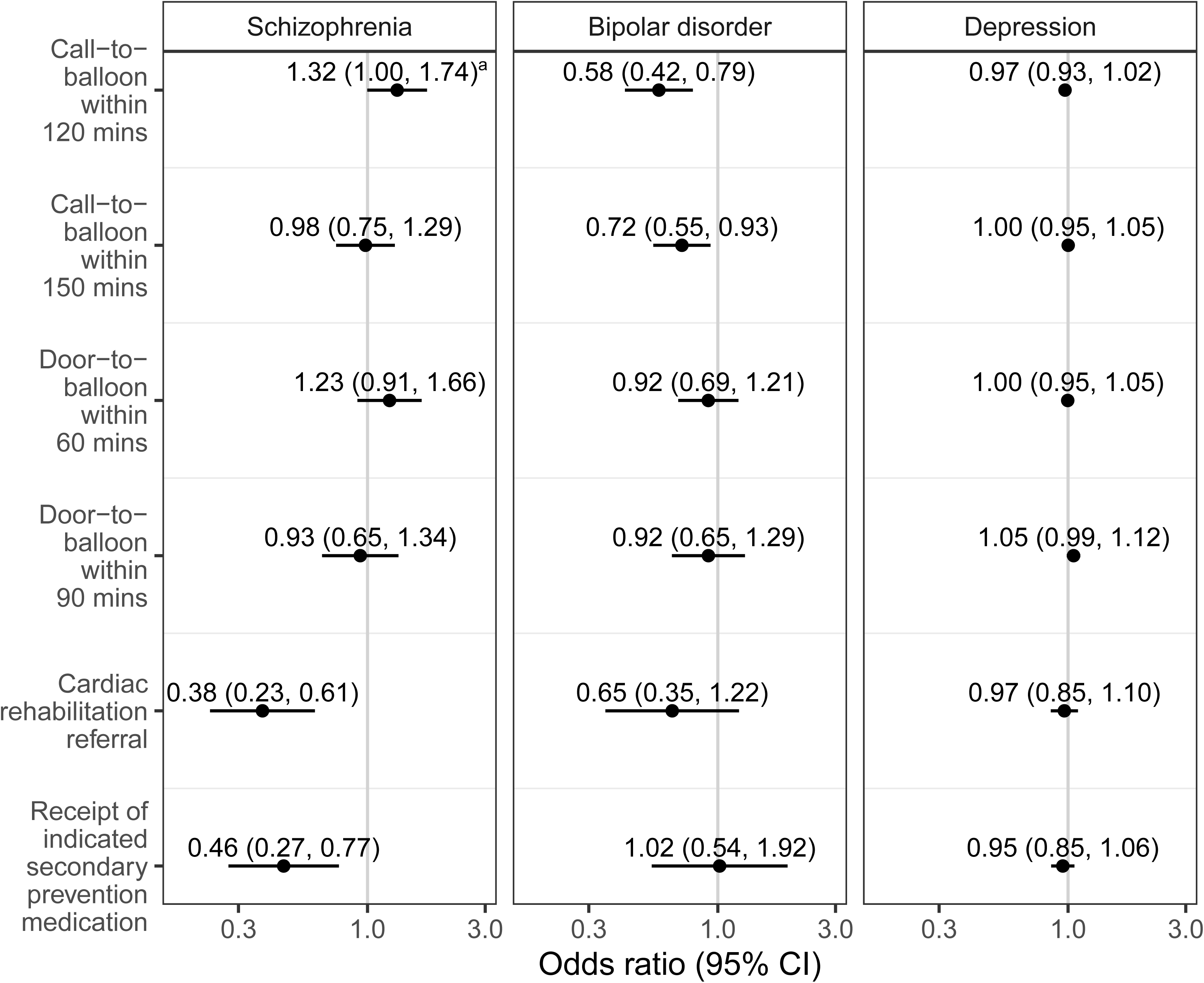
Odds ratios for receipt of each care standard following STEMI in patients with schizophrenia, bipolar disorder or depression versus those without any of these disorders. Odds ratios are adjusted for sociodemographic characteristics, hospital, MI timing, comorbidities and MI presentation features (model 5).

### Sensitivity analyses

Findings from models that included patients with complete information on sociodemographic, timing and clinical history variables, but did not adjust for MI presentation features, were similar to those obtained in the primary analysis, for both NSTEMI and STEMI (Supplementary data online, *Figures S4 and S5*). Based on the fully adjusted models, the sensitivity analyses identified the same statistically significant disparities that were identified by the primary analyses. Additionally, for NSTEMI, the sensitivity analyses also identified an association between schizophrenia and receipt of indicated secondary prevention medication (OR: 0.72 (0.56, 0.93)).

### COVID-19 and mental disorder differences in receipt of care

Within the study period, overall monthly admissions for both NSTEMI and STEMI were lowest at the start of the COVID-19 pandemic, in March and April 2020 (Supplementary data online, *Figure S6*). For NSTEMI, the proportions of patients considered eligible for angiography and receiving angiography were lowest, whilst the proportion receiving angiography within 72 hours was highest, in April 2020. Following NSTEMI, the proportions of patients admitted to a cardiac ward, referred for rehabilitation and receiving indicated secondary prevention medication were relatively stable over the study period. For STEMI, the proportion of patients in whom CTB and DTB time targets were met decreased over the study period, whereas the proportions of patients referred for rehabilitation and receiving appropriate secondary prevention medication were relatively stable. Time trends were generally similar for patients with versus without schizophrenia, bipolar disorder or depression (Supplementary data online, *Figure S7*).

For both NSTEMI and STEMI, there was no evidence that the associations between mental disorder and receipt of care varied over our study period, with one exception (Supplementary data online, *Table S3*). For STEMI, the association between mental disorder and CTB within 120 minutes varied across the study period. There was a statistically significant difference between the association in the pre-pandemic period (November 2019 – February 2020), and the association in the period July 2021 – October 2021; however, neither association indicated a difference between people with vs without schizophrenia, bipolar disorder or depression (Supplementary data online, *Table S4*). For the sensitivity analysis, for both NSTEMI and STEMI, there was no evidence that the associations between mental disorder and receipt of care varied over our study period (Supplementary data online, *Table S5*).

## Discussion

Among people with NSTEMI, a history of schizophrenia, bipolar disorder or depression was associated with lower odds of angiography eligibility and receipt (if eligible), cardiac ward admission and cardiac rehabilitation referral, with the largest disparities for people with schizophrenia.

Depression was additionally associated with lower receipt of secondary prevention medication. In people with a STEMI, those with bipolar disorder were less likely to meet targets for call-to-balloon time and people with schizophrenia had lower odds of cardiac rehabilitation referral and receipt of indicated secondary prevention medication. There were no differences in receipt of clinical care standards for STEMI between people with depression and the comparison group. Within our study period, which included the acute phase of the COVID-19 pandemic and its aftermath, receipt of most clinical care standards was stable over time. However, for NSTEMI, receipt of angiography within 72 hours peaked in April 2020, when admissions were at their lowest, and for STEMI the proportion of patients in whom time to balloon targets were met decreased over the study period. Trends over time were similar for people with and without a mental disorder, hence disparities in care were largely unaffected by the COVID-19 pandemic.

### Disparities by mental disorder

Our findings on mental disorder and receipt of care align with and substantially add to those of existing studies. Whilst there are consistent reports from other universal/near-universal healthcare settings of lower receipt of angiography^31–35^ and revascularisation procedures^11,31–36^ in people with mental disorders, previous studies largely focused on psychoses.^31,32,34,36^ Our study makes a novel contribution by reporting on a wider set of guideline-indicated care standards and including mood disorders in addition to schizophrenia. To our knowledge, it is the first to report on receipt of these care standards by mental disorder status. Our separate analysis of patients with NSTEMI and STEMI also provides additional insight, revealing differing patterns of disparities by MI type and mental disorder.

Our study reveals lower rates of interventional treatment for NSTEMI in people with not only schizophrenia, but also bipolar disorder and depression. These disparities are not as evident STEMI treatment. This is perhaps unsurprising given that NSTEMI management has less of a focus on rapidity of intervention and a more staged approach to assessment, investigation, risk assessment and clinical management decisions, with more opportunity for inequities to occur in the care pathway. The NSTEMI disparities in angiography and PCI may partly be influenced by the higher prevalence of comorbidities, particularly diabetes, among patients with mental disorder. People with diabetes experience disparities in receipt of hospital care for MI, including reduced likelihood of receipt of PCI.^37^ However, the persistence of associations following adjustment for comorbidities suggest other factors also play a role. The observed reduced likelihood of admission to a cardiac ward may contribute. This suggests that people with a mental disorder have reduced access to dedicated, concentrated specialist nursing and medical care. Patients with NSTEMI admitted to a cardiac ward are more likely to receive guideline directed management, including angiography, PCI and optimal secondary prevention medication on discharge.^38^ Moreover, cardiac ward admission is linked to long-term prognostic benefits, such as improved medication titration, early cardiac rehabilitation referral, and structured discharge planning, which are thus disproportionately withheld from people with a mental disorder. We did not have information on reason for non-admission to a cardiac ward, but this may relate to delayed diagnosis among people with a mental disorder, potentially due to atypical symptom presentation or ‘diagnostic overshadowing’, whereby physical health symptoms are misattributed to mental health conditions.^39^

It is reassuring that, where PCI is conducted, DTB targets, a marker of hospital performance, didn’t differ by mental disorder. The lower achievement of CTB targets in people with bipolar disorder, but not schizophrenia or depression, is intriguing, and implies that this sub-population experiences delays in pre-hospital care, the reasons for which should be investigated further.

Reasons for mental disorder disparities in acute MI care have been little investigated in previous studies. However, our recent qualitative study included interviews with health care professionals involved in MI care provision across the acute care pathway^40^ and people with a mental disorder who had experienced an MI *(unpublished data)*. We identified multiple practitioner-, patient- and system-related challenges that may impact delivery of optimal MI care for people with a mental disorder. These included: stigma towards patients with a mental disorder and lack of confidence in managing patient behaviour, inadequate mental health training, under-staffing, difficulties accessing input from psychiatric team colleagues, poor patient-practitioner engagement and communication, diminished patient history capacities, and intervention and medication compliance concerns.

Beyond the acute treatment phase of the clinical care pathway, our study adds to existing evidence of lower secondary prevention medication prescribing post-MI in people with a mental disorder from universal healthcare settings^31,32,34,35^ by demonstrating lower likelihood of such prescribing at point of discharge for people with schizophrenia (following STEMI) and depression (following NSTEMI) in England. Some medications, such as beta-blockers may be avoided due to perceived risks (for example QT-interval prolongation^41^). However, our findings support the need for structured documentation of contraindications to ensure optimal prescribing of vital secondary preventive medication. There is also a need to challenge assumptions that people with a mental disorder or those with a history of poor adherence to psychotropic medication are less likely to adhere to cardiovascular medication. Indeed, people with mental disorder may be more likely to adhere to cardiovascular medication.^42^ To our knowledge, no previous study has reported on referral for, or receipt of, cardiac rehabilitation by mental disorder. Reduced participation in cardiac rehabilitation has been linked to various factors, including low socioeconomic status, smoking and history of coronary heart disease,^43^ all of which are more common in people with a mental disorder. This may lead to erroneous assumptions around cardiac rehabilitation on the part of healthcare professionals influencing referral decisions.

### Impact of the COVID-19 pandemic

Whilst the observed mental disorder disparities in guideline-informed care indicators are concerning, it is reassuring that the COVID-19 pandemic did not worsen or create new disparities in MI care. This reflects other work on the impact of the COVID-19 pandemic on mental disorder disparities, which found no evidence that disparities in CVD incidence in England were worsened during or following the pandemic.^44^ The overall reduction in angiography and PCI in the whole study population during the first lockdown aligns with the well-documented reduction in MI hospitalisation during that time^45^, with reduced patient burden perhaps accounting for the greater proportion with NSTEMI receiving angiogram within 72 hours. For STEMI, we observed a deterioration in the time to balloon targets over our study period, but reports from MINAP suggest this is part of a longer-term trend, which predates the pandemic.^46^

### Implications

Whilst our study adds greater breadth and depth to current understanding of mental disorder disparities in acute cardiovascular care, it does raise further questions to be addressed in future research. Our qualitative study (publication forthcoming) provides much-needed insight to the experiences of and challenges faced by patients and health care professionals, but further granular quantitative *and* qualitative research is needed. Quantitative studies should aim to: explore detailed differences by mental disorder in the in-hospital patient journey (considering factors such as symptom presentation, time to diagnosis, ward transitions and angiogram findings); identify factors associated with sub-optimal prescribing at point of discharge; and investigate access to and initiation of cardiac rehabilitation. Future research should also examine the extent to which these disparities in receipt of guideline-informed care standards account for mental disorder disparities in MI outcomes in this population.

In the meantime, there are implications for improving the delivery and clinical audit of acute MI care for people with a mental disorder. First, greater recognition of mental disorder disparities in cardiovascular care is desperately needed. A recent Lancet series on variations in cardiovascular disease outcomes included very little focus on mental disorder disparities in cardiovascular disease^47^, being notably absent from the synthesis of inequalities in MI healthcare delivery and outcomes from the UK.^48^ In addition to age, sex, deprivation and ethnicity, we recommend that mental disorder, often neglected, be included alongside more established sociodemographic drivers of healthcare inequalities. Mental disorder should be included as a stratifying variable in national cardiovascular audits and quality improvement dashboards, similar to ethnicity or deprivation status. Second, approaches to addressing impediments to delivery of optimal guideline-informed care are urgently needed. These include for example improved integrated care through enhanced collaboration between cardiologists and liaison psychiatrists to effectively manage any potentially disruptive mood or behavioural symptoms that might negatively impact delivery of care and to provide expert psychotropic medication management advice. Service innovations are urgently needed to address disparities in cardiac rehabilitation access and could include co-designed, tailored cardiac rehabilitation pathways that integrate psychiatric input, flexible schedule and peer support models.

### Strengths and limitations

Our study has various strengths. To our knowledge it is the first study to comprehensively investigate receipt of clinical guideline-informed acute care for MI by mental disorder status and the impact of the COVID-19 pandemic on mental disorder disparities. The use of a national and representative MI audit along with objective ascertainment of mental disorder from primary care records minimises both selection and information bias and provides good generalisability. The large study population allowed for separate analysis of NSTEMI and STEMI, adding valuable insights by MI type, which has rarely been addressed previously. We were also able to take account of sociodemographic factors, hospital admission factors, clinical history and MI presentation characteristics in analyses. Finally, the use of data from a contemporary time period is an important strength, given the evolution of clinical guidelines for acute cardiac care and increase in access to revascularisation procedures over time.

Our study has some limitations. Our analyses focused on MI care in specific mental disorders and findings may not be generalisable to other mental disorders. Also, since we did not exclude other mental disorders from the comparison group, effect estimates may be underestimated if similar disparities are observed for people with other mental disorders. Most depression codes used in primary care do not reflect severity. Whilst we had planned to focus on severe mental illness (schizophrenia, bipolar disorder and major depression), in order to evaluate disparities in people with the most severe mental disorders, due to the lack of specificity in the depression codes we widened our definition of depression to include depression of any severity. Since we were unable to investigate receipt of care by severity of depression, our findings for depression may not be generalisable to people with severe depression, in whom disparities may be even larger. Although care data was largely complete, a higher proportion of patients had missing data for the time to treatment outcomes. Similarly, whilst covariate data were mostly nearly complete, missingness was higher for MI presentation characteristics and generally more likely to be missing for people with schizophrenia. Reassuringly, findings were robust to analyses adjusting for sociodemographic variables, timing, and clinical history, but not MI presentation characteristics. We performed complete case analysis rather than conducting multiple imputation given that for each outcome covariates were complete for at least 80% of participants with outcome data in the primary analyses and 97% of participants with outcome data in the sensitivity analyses where we excluded the MI presentation characteristics. Our analysis of the impact of COVID-19 on mental disorder-receipt of care associations was limited by complete primary care data only being available from November 2019. With only four months of pre-pandemic data, we could not robustly account for seasonal fluctuations in receipt of care. Finally, lower numbers of people with schizophrenia and bipolar disorder precluded investigation of interactions between individual mental disorders and time period for receipt of care; our analyses may not have identified any nuanced impact of the pandemic on quality of care for the less common conditions of schizophrenia and bipolar disorder.

## Conclusions

Our study found that people with schizophrenia, bipolar disorder or depression are less likely to receive guideline-informed care for MI than people without any of these disorders. Disparities were consistently observed across almost all NSTEMI care standards, for schizophrenia, bipolar disorder and depression, with findings more mixed, but with generally fewer disparities, for STEMI. Health care professionals involved in the delivery of hospital care for MI should be better supported to deliver optimal care to patients with mental health comorbidities across the entire acute cardiac care pathway, from symptom assessment through to referral for cardiac rehabilitation.

## Author contributions

C.J. conceived the study. C.J., K.F., D.C., S.W.M, S.P and D.J.S. were awarded funding for the study. J.N. extracted the analysis dataset. K.F. conducted the statistical analysis. All authors contributed to the interpretation of the results. K.F. and C.J. wrote the first draft of the article. All authors commented on the draft and approved the final version.

## Data Availability

The protocol, code lists and data curation and analysis code for this study are available on GitHub (https://github.com/BHFDSC/CCU046_01).

The data used in this study are available in NHS England’s Secure Data Environment (SDE) service for England, but as restrictions apply they are not publicly available (https://digital.nhs.uk/services/secure-data-environment-service). The CVD-COVID-UK/COVID-IMPACT programme, led by the BHF Data Science Centre (https://bhfdatasciencecentre.org/), received approval to access data in NHS England’s SDE service for England from the Independent Group Advising on the Release of Data (IGARD) (https://digital.nhs.uk/about-nhs-digital/corporate-information-and-documents/independent-group-advising-on-the-release-of-data) via an application made in the Data Access Request Service (DARS) Online system (ref. DARS-NIC-381078-Y9C5K) (https://digital.nhs.uk/services/data-access-request-service-dars/dars-products-and-services). The CVD-COVID-UK/COVID-IMPACT Approvals & Oversight Board (https://bhfdatasciencecentre.org/areas/cvd-covid-uk-covid-impact/) subsequently granted approval to this project to access the data within NHS England’s SDE service for England. The de-identified data used in this study were made available to accredited researchers only. Those wishing to gain access to the data should contact in the first instance.

## Acknowledgements

This work was carried out with the support of the BHF Data Science Centre led by HDR UK (BHF Grant no. SP/19/3/34678). This study made use of de-identified data held in NHS England’s Secure Data Environment service for England and made available via the BHF Data Science Centre’s CVD-COVID-UK/COVID-IMPACT consortium. The BHF DSC Health Data Science Team provided data curation resources and support. Members of the CVD-COVID-UK/COVID-IMPACT Consortium provided feedback on a draft of the manuscript. This work used data provided by patients and collected by the NHS as part of their care and support. We would also like to acknowledge all data providers who make health relevant data available for research.

## Funding

This study was funded by a Chief Scientist Office Scotland grant (Ref HIPS/21/48) awarded to C.J., K.F., D.C., D.J.S., S.W.M. and S.P. The authors K.F., D.J.S. and C.J. are also supported by the UKRI-funded University of Edinburgh Hub for Metabolic Psychiatry (Ref MR/Z503563/1), within the UK Mental Health Platform (Ref MR/Z000548/1). The British Heart Foundation Data Science Centre (grant No SP/19/3/34678, awarded to Health Data Research (HDR) UK) funded co-development (with NHS England) of the Secure Data Environment service for England, provision of linked datasets, data access, user software licences, computational usage, and data management and wrangling support, with additional contributions from the HDR UK Data and Connectivity component of the UK Government Chief Scientific Adviser’s National Core Studies programme to coordinate national COVID-19 priority research. Consortium partner organisations funded the time of contributing data analysts, biostatisticians, epidemiologists, and clinicians. RS is part-funded by: i) the NIHR Maudsley Biomedical Research Centre at the South London and Maudsley NHS Foundation Trust and King’s College London; ii) the National Institute for Health Research (NIHR) Applied Research Collaboration South London (NIHR ARC South London) at King’s College Hospital NHS Foundation Trust; iii) UKRI – Medical Research Council through the DATAMIND HDR UK Mental Health Data Hub (MRC reference: MR/W014386); iv) the UK Prevention Research Partnership (Violence, Health and Society; MR-VO49879/1), an initiative funded by UK Research and Innovation Councils, the Department of Health and Social Care (England) and the UK devolved administrations, and leading health research charities.

## Disclosure of interest

K.F., J.N., D.C., S.W.M., S.P., D.J.S., R.S., A.V. and C.A.J. declare no disclosures of interest for this work. C.B. is employed by the University of Glasgow which holds consultancy and research agreements for his work with Abbott Vascular, AstraZeneca, Boehringer Ingelheim, CorFlow, MSD, Novartis, Servier, Siemens Healthcare, Xylocor and Zoll Medical.

## Ethical approval

The North East - Newcastle and North Tyneside 2 research ethics committee provided ethical approval for the CVD-COVID-UK/COVID-IMPACT research programme (REC No 20/NE/0161) to access, within secure trusted research environments, unconsented, whole-population, de-identified data from electronic health records collected as part of patients’ routine healthcare.

## Supplementary Material

### Methods S1: SNOMED code lists for mental disorders

The General Practice Extraction Service Data for Pandemic Planning and Research (GDPPR) dataset includes a subset of all SNOMED codes used in the United Kingdom. The subset of codes included is defined by the primary care domain (PCD) reference set.^1^ The PCD reference set is also used by the Quality and Outcomes Framework (QOF) and current and historical versions of the reference set are available from NHS England.^2^ The available SNOMED codes are grouped into code clusters.^1^

To develop our SNOMED code lists for mental disorders we extracted codes from all of the clusters related to relevant mental disorder diagnoses: DEPR_COD (depression diagnosis codes), DEPRES_COD (depression resolved codes) and MH_COD (psychosis and schizophrenia and bipolar affective disease codes) from version 47.1 of the QOF cluster list (which was the most recent version at the time of developing the SMI code lists).^3^ Additionally, we mapped existing Read v2 and Clinical Terms Version 3 code lists^4^ for schizophrenia, bipolar disorder and depression to SNOMED, and identified codes available within the GDPPR dataset. We collated all SNOMED codes identified from the clusters, and from the mapping process and then identified codes for each of schizophrenia, bipolar disorder or depression.

All of our SNOMED code lists are available via the HDR UK Phenotype Library:

- **Schizophrenia:** https://phenotypes.healthdatagateway.org/phenotypes/PH1718
- **Bipolar disorder:** https://phenotypes.healthdatagateway.org/phenotypes/PH1719
- **Depression:** https://phenotypes.healthdatagateway.org/phenotypes/PH1720

## Methods S2: Definition of care outcomes

| MI Type | Outcome | Included patients | Outcome measure |
| --- | --- | --- | --- |
| NSTEMI | Eligibility for angiography | All | Patients considered eligible for angiography (MINAP coronary angiography field [4.13] indicates that angiography was performed or is planned after discharge, that angiography was not performed or that the patient refused) |
|  | Receipt of angiography | Patients considered eligible for angiography, as defined above | Patient received angiography (MINAP coronary angiography field [4.13] indicates that angiography was performed) |
|  | Angiography within 72 hours | Patients who received angiography, as defined above | Angiography within 72 hours, where time to angiography is calculated as the number of hours from arrival at hospital (MINAP field 3.06) to local angiography (MINAP field 4.18) |
|  | Admission to cardiac ward | Patients who did not die in A&E (admission ward [MINAP field 3.17] is not recorded as 'Died in A&E') | Following the definition of this target in the MINAP 2022 annual report<sup>1</sup>, patients where admission ward (MINAP field 3.17) is recorded as 'cardiac care unit', 'intensive therapy unit' or 'cardiac ward (non CCU)' |
| STEMI | Call-to-balloon time within 120 minutes | Patients who underwent primary PCI. Initial reperfusion treatment (MINAP field 3.39) recorded as 'pPCI in house', 'pPCI already was performed at the interventional hospital' or 'Referred for consideration for pPCI elsewhere' | Call-to-balloon within 120 minutes, where call-to-balloon time is calculated as the number of minutes from the call for help (MINAP field 3.02) to reperfusion treatment (MINAP field 3.09) |
|  | Call-to-balloon time within 150 minutes |  | Call-to-balloon within 150 minutes, where call-to-balloon time is calculated as described above |
|  | Door-to-balloon time within 60 minutes |  | Door-to-balloon within 60 minutes, where door-to-balloon time is calculated as the number of minutes from arrival at hospital (MINAP field 3.06) to reperfusion treatment (MINAP field 3.09) |
|  | Door-to-balloon time within 90 minutes |  | Door-to-balloon within 90 minutes, where door-to- balloon time is calculated as described above |
| NSTEMI & STEMI | Suitable secondary prevention medication prescribed at discharge | Patients who were discharged home. Discharge destination (MINAP field 4.16) recorded as 'Home' | Where indicated, patient prescribed (or refused) aspirin (MINAP field 4.08), beta blocker (MINAP field 4.05), statin (MINAP field 4.07), either ACE inhibitor or angiotensin receptor antagonist (MINAP field 4.06), and either thienopyridine (MINAP field 4.27) or ticagrelor (MINAP field 4.31). For each field, we defined that the patient had prescribed (or refused) the medication where indicated if the response was recorded as 'Yes', 'Contraindicated', 'Patient declined treatment', 'Not applicable' or 'Not indicated'. |
|  | Referral for cardiac rehabilitation | Patients who were discharged home and did not have referral for cardiac rehabilitation (MINAP field 4.09) recorded as 'Not indicated' (the MINAP specification indicates that cardiac rehabilitation may not be indicated due to severe co-morbidity, for example). | Referral for cardiac rehabilitation (MINAP field 4.09) recorded as 'Yes'. |

## Methods S3: Additional description of covariates

### Admission timing

We used the MINAP hospital arrival time variable to determine whether the admission was during the day (6am – 6pm) or overnight.

### Admission day

We used the MINAP hospital arrival time variable to determine whether the admission was on a weekday or weekend.

### 4-month period of admission

We used the MINAP hospital arrival time variable to identify the 4-month period during which the MI admission occurred, starting from the pre-pandemic period of November 2019 to February 2020.

### Hospital

We ascertained the hospital the patient was admitted to by linking the MINAP record to HES APC records.

### Comorbid angina

We identified comorbid angina if any of the following occurred:

- The MINAP previous angina field (field 2.06) recorded “1. Yes”
- The patient had a diagnosis of angina recorded in GDPPR prior to their date of MI admission.
- The patient had a diagnosis of angina recorded in HES APC prior to their date of MI admission.

We identified diagnoses using existing SNOMED (GDPPR) and ICD-10 (HES APC) code lists for angina which are available on the HDR UK Phenotype Library.^1^

### Comorbid cerebrovascular disease

We identified comorbid cerebrovascular disease if any of the following occurred:

- The MINAP cerebrovascular disease field (field 2.10) recorded “1. Yes”
- The patient had a diagnosis of cerebrovascular disease recorded in GDPPR prior to their date of MI admission. We identified diagnoses using SNOMED codes in the haemorrhagic stroke (HSTRK_COD), non-haemorrhagic stroke (OSTR_COD), stroke diagnosis (STRK_COD) and transient ischaemic attack (TIA_COD) code clusters (see Methods S2 for further explanation of code clusters) and additionally the SNOMED codes 195163003, 67992007 and 73192008.
- The patient had a diagnosis of cerebrovascular disease recorded in HES APC prior to their date of MI admission. We identified diagnoses using ICD-10 codes I60-I69 and G45.

Our code lists for comorbid cerebrovascular disease are available on our GitHub repository (https://github.com/BHFDSC/CCU046_01) and via the HDR UK Phenotype Library: https://phenotypes.healthdatagateway.org/phenotypes/PH1791.

### Comorbid chronic renal failure

We identified comorbid chronic renal failure if any of the following occurred:

- The MINAP chronic renal failure field (field 2.12) recorded “1. Yes”
- The patient had a diagnosis of chronic renal failure recorded in GDPPR prior to their date of MI admission. We identified diagnoses using SNOMED codes. We developed the code list by identifying chronic renal failure codes from an existing chronic kidney disease code list^2^ and the GDPPR code cluster for chronic kidney disease stage 4 and 5 (CKDATRISK1_COD).
- The patient had a diagnosis of chronic renal failure recorded in HES APC prior to their date of MI admission. We identified diagnoses using ICD-10 codes. We developed the code list by identifying chronic renal failure codes from an existing chronic kidney disease code list^2^ and existing end stage renal disease code lists.^3,4^

Our code lists for comorbid chronic renal failure are available on our GitHub repository (https://github.com/BHFDSC/CCU046_01) and via the HDR UK Phenotype Library: https://phenotypes.healthdatagateway.org/phenotypes/PH1792.

### Comorbid diabetes

We identified comorbid diabetes if any of the following occurred:

- The MINAP diabetes field (field 2.17) recorded “1. Diabetes (dietary control)” or “2. Diabetes (oral medicine)” or “3. Diabetes (insulin)” or “5. Insulin plus oral medication”
- The patient had a diagnosis of diabetes recorded in GDPPR prior to their date of MI admission.
- The patient had a diagnosis of diabetes recorded in HES APC prior to their date of MI admission.

We identified diagnoses using existing SNOMED (GDPPR) and ICD-10 (HES APC) code lists for diabetes which are available on the HDR UK Phenotype Library.^5^

### Comorbid heart failure

We identified comorbid heart failure if any of the following occurred:

- The MINAP heart failure field (field 2.13) recorded “1. Yes”
- The patient had a diagnosis of heart failure recorded in GDPPR prior to their date of MI admission.
- The patient had a diagnosis of heart failure recorded in HES APC prior to their date of MI admission.

We identified diagnoses using existing SNOMED (GDPPR) and ICD-10 (HES APC) code lists for heart failure which are available on the HDR UK Phenotype Library.^6^

### Previous MI

We identified previous MI if either of the following occurred:

- The patient had a diagnosis of MI recorded in GDPPR more than 3 days prior to their date of MI admission (to ensure that we didn’t count the index event). We identified diagnoses by adapting an existing MI SNOMED code list.^7^
- The patient had a diagnosis of MI recorded in HES APC more than 3 days prior to their date of MI admission. We identified diagnoses using ICD-10 codes (I21, I22) as per previous studies from the authors.^8^

Our code lists for myocardial infarction are available on our GitHub repository (https://github.com/BHFDSC/CCU046_01) and via the HDR UK Phenotype Library (https://phenotypes.healthdatagateway.org/phenotypes/PH1722).

We couldn’t use the MINAP previous MI field (field 2.05) because it wasn’t included in the version of the MINAP dataset held by the CVD-COVID-UK/COVID-IMPACT Consortium at the time of data curation.

### Previous PCI

We identified previous PCI if any of the following occurred:

- The MINAP previous PCI field (field 2.18) recorded “1. Yes”
- The patient had a PCI recorded in HES APC prior to their date of MI admission. We identified PCI using the OPCS-4 procedure code K75, in line with previous studies from the authors.^8^

We didn’t use GDPPR to identify PCI because the GDPPR clusters do not include a comprehensive set of PCI codes.

### COVID-19 positive

We identified patients who had COVID-19 within 14 days either side of their MI admission based on diagnoses recorded in GDPPR or HES APC, hospitalised cases of COVID-19 recorded in COVID-19 Hospitalisations in England Surveillance System (CHESS) and positive tests for COVID-19 recorded in Second Generation Surveillance System (SGSS) or Secondary Uses Service (SUS).

---

**Figure S1:**
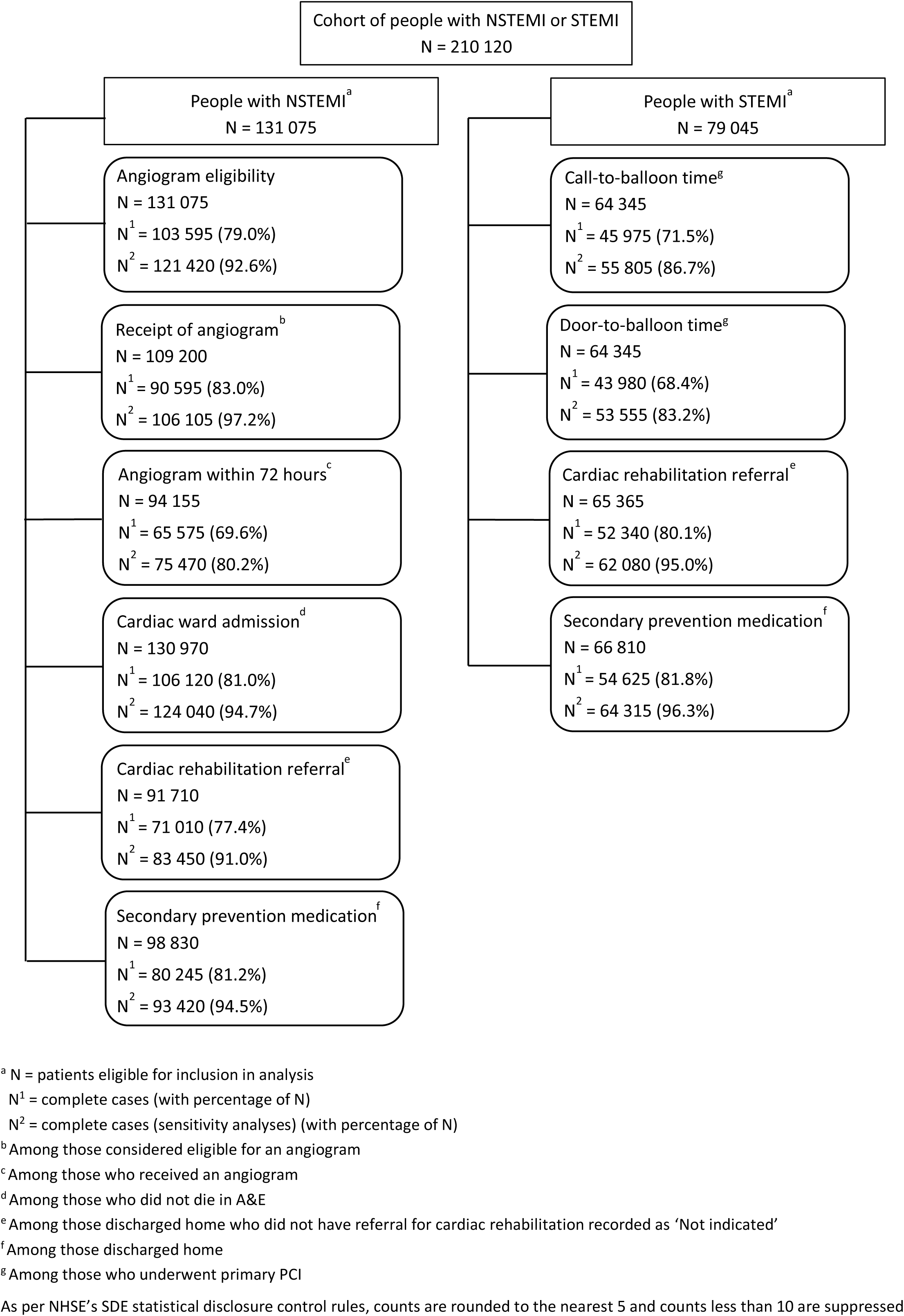
Flow diagram for the analysis cohorts

**Figure S2:**
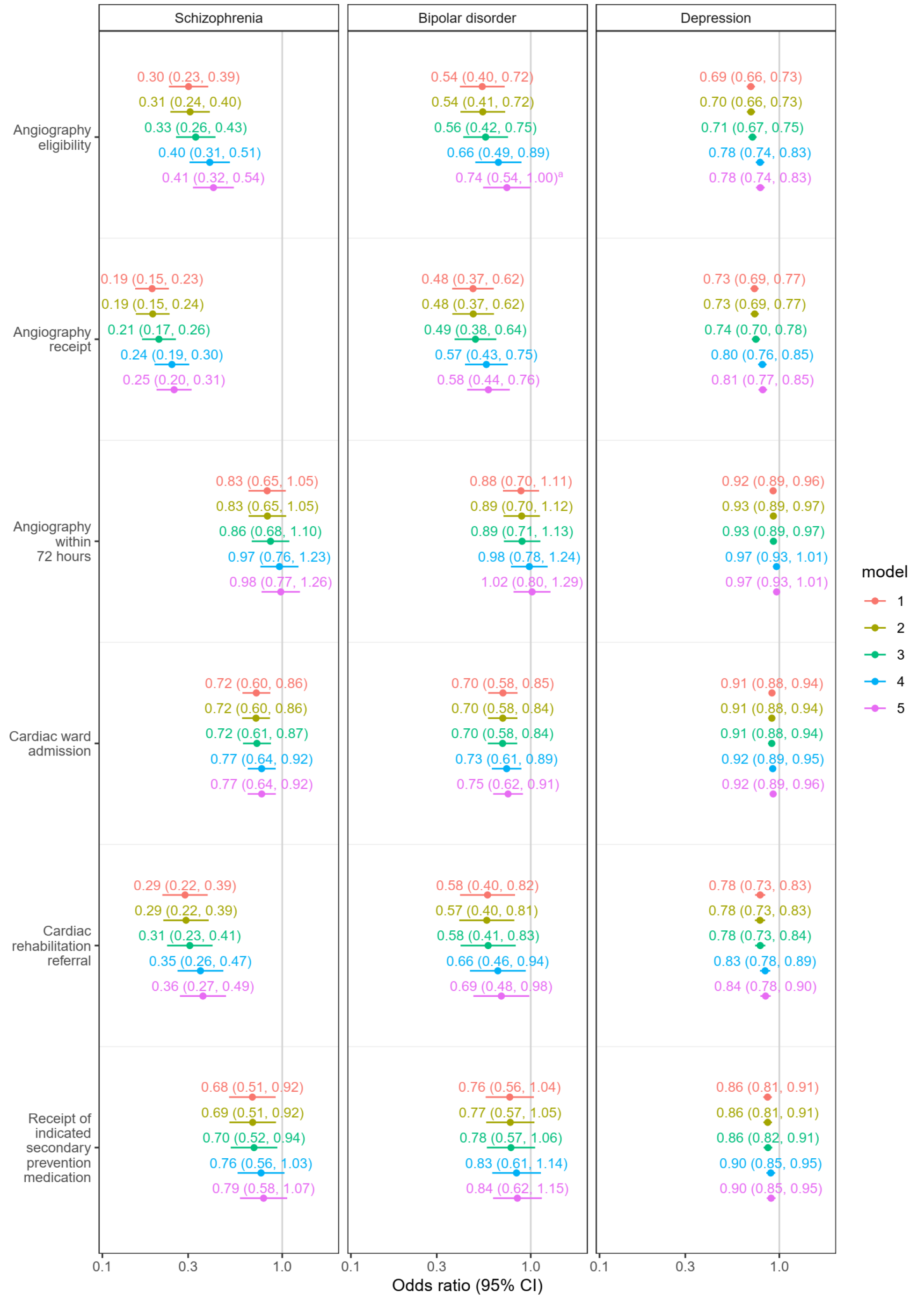
Odds ratios for receipt of each care standard following NSTEMI in patients with schizophrenia, bipolar disorder or depression versus those without any of these disorders. Models 1 to 5. Model 1 adjusts for age, sex and hospital (random effect); model 2 additionally adjusts for timing variables; model 3 adds ethnicity and area-level deprivation; model 4 adds clinical history; and model 5 adds MI presentation features. ^a^ 0.736 (0.543, 0.998)

**Figure S3:**
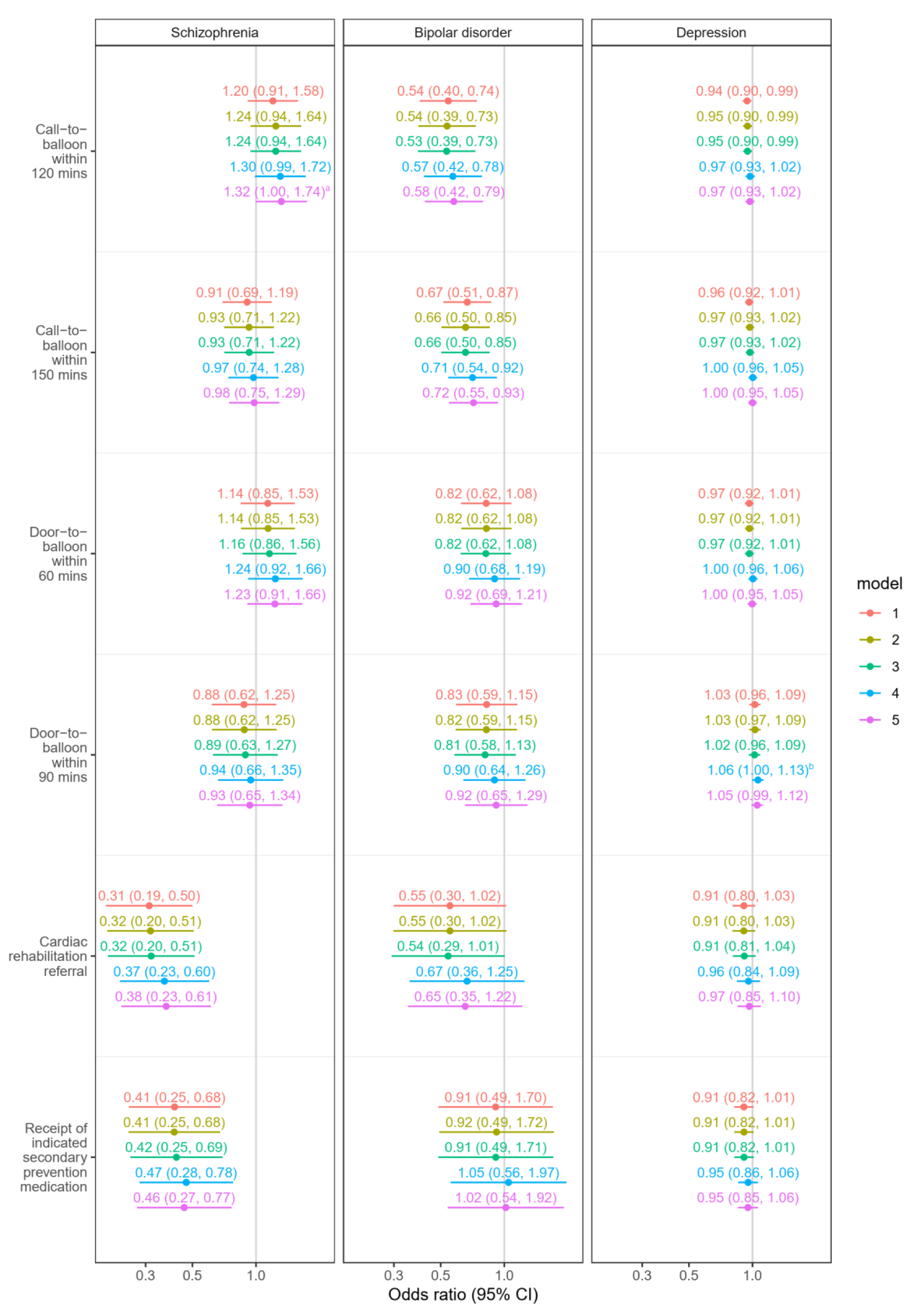
Odds ratios for receipt of each care standard following STEMI in patients with schizophrenia, bipolar disorder or depression versus those without any of these disorders. Models 1 to 5. Model 1 adjusts for age, sex and hospital (random effect); model 2 additionally adjusts for timing variables; model 3 adds ethnicity and area-level deprivation; model 4 adds clinical history; and model 5 adds MI presentation features. ^a^ 1.319 (0.998, 1.744) ^b^ 1.061 (0.997, 1.130)

**Figure S4:**
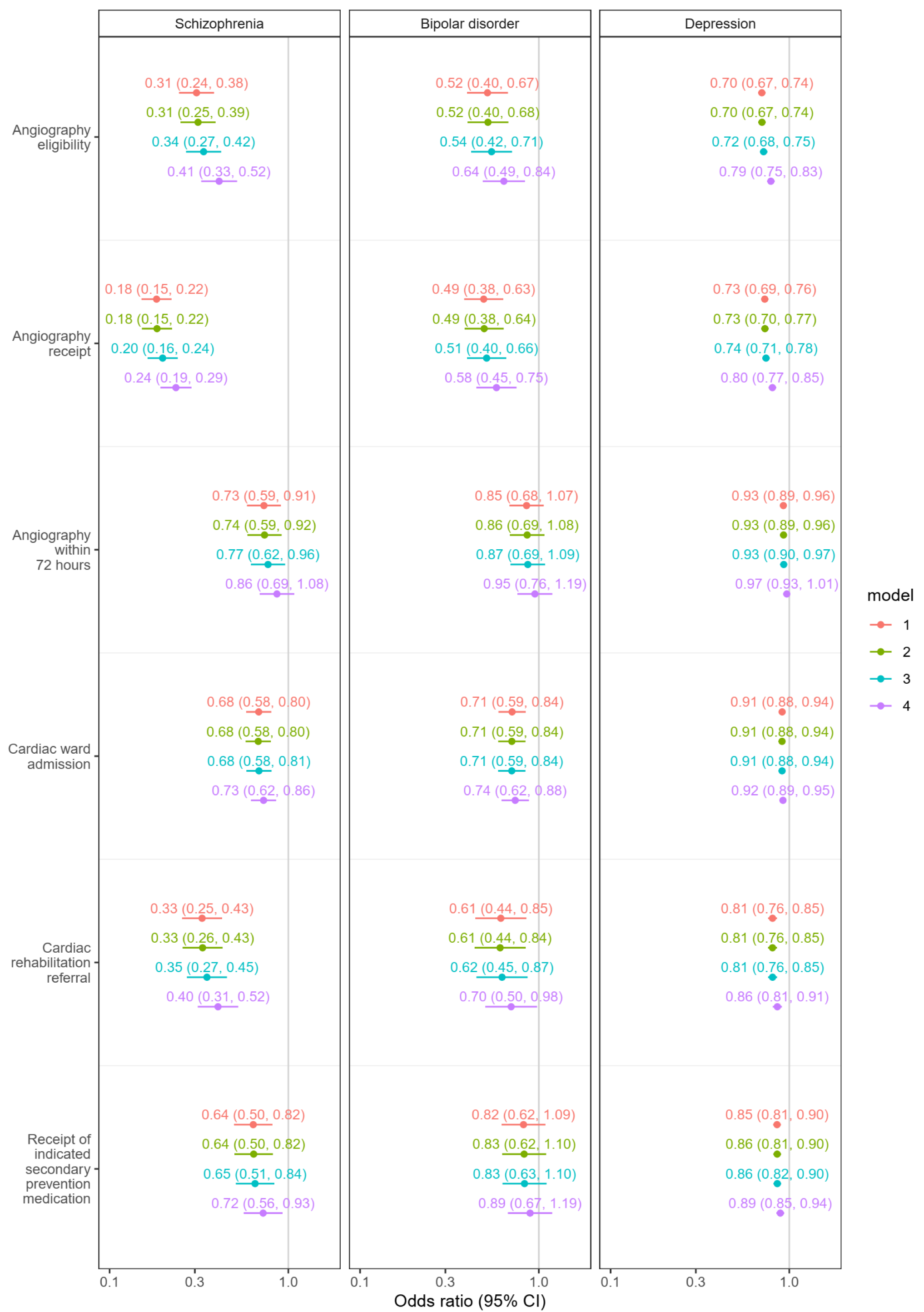
Odds ratios for receipt of each care standard following NSTEMI in patients with schizophrenia, bipolar disorder or depression versus those without any of these disorders. Sensitivity analysis. Models 1 to 4. Model 1 adjusts for age, sex and hospital (random effect); model 2 additionally adjusts for timing variables; model 3 adds ethnicity and area-level deprivation; and model 4 adds clinical history.

**Figure S5:**
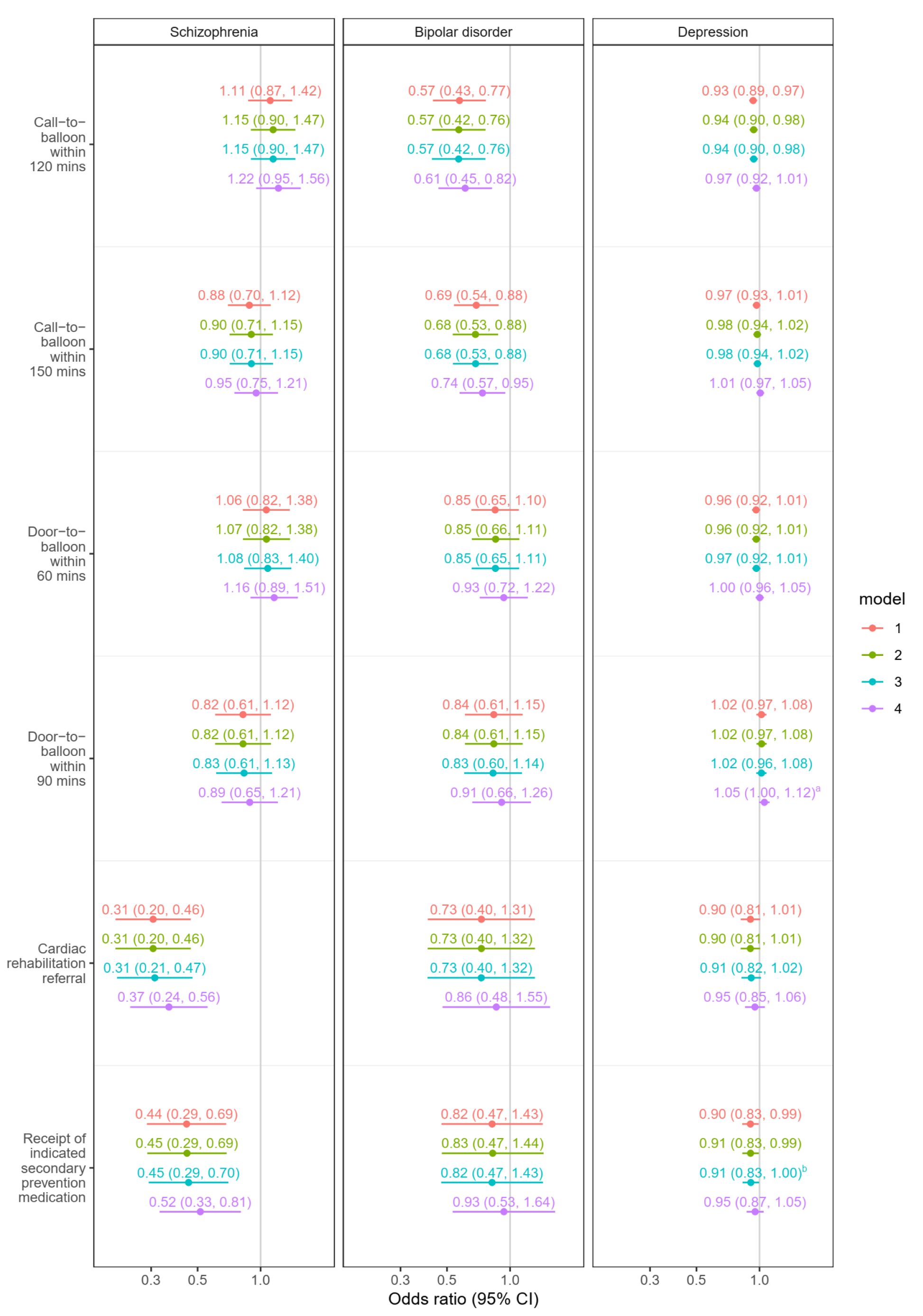
Odds ratios for receipt of each care standard following STEMI in patients with schizophrenia, bipolar disorder or depression versus those without any of these disorders. Sensitivity analysis. Models 1 to 4. Model 1 adjusts for age, sex and hospital (random effect); model 2 additionally adjusts for timing variables; model 3 adds ethnicity and area-level deprivation; and model 4 adds clinical history. ^a^ 1.055 (0.996, 1.117) ^b^ 0.909 (0.829, 0.996)

**Figure S6:**
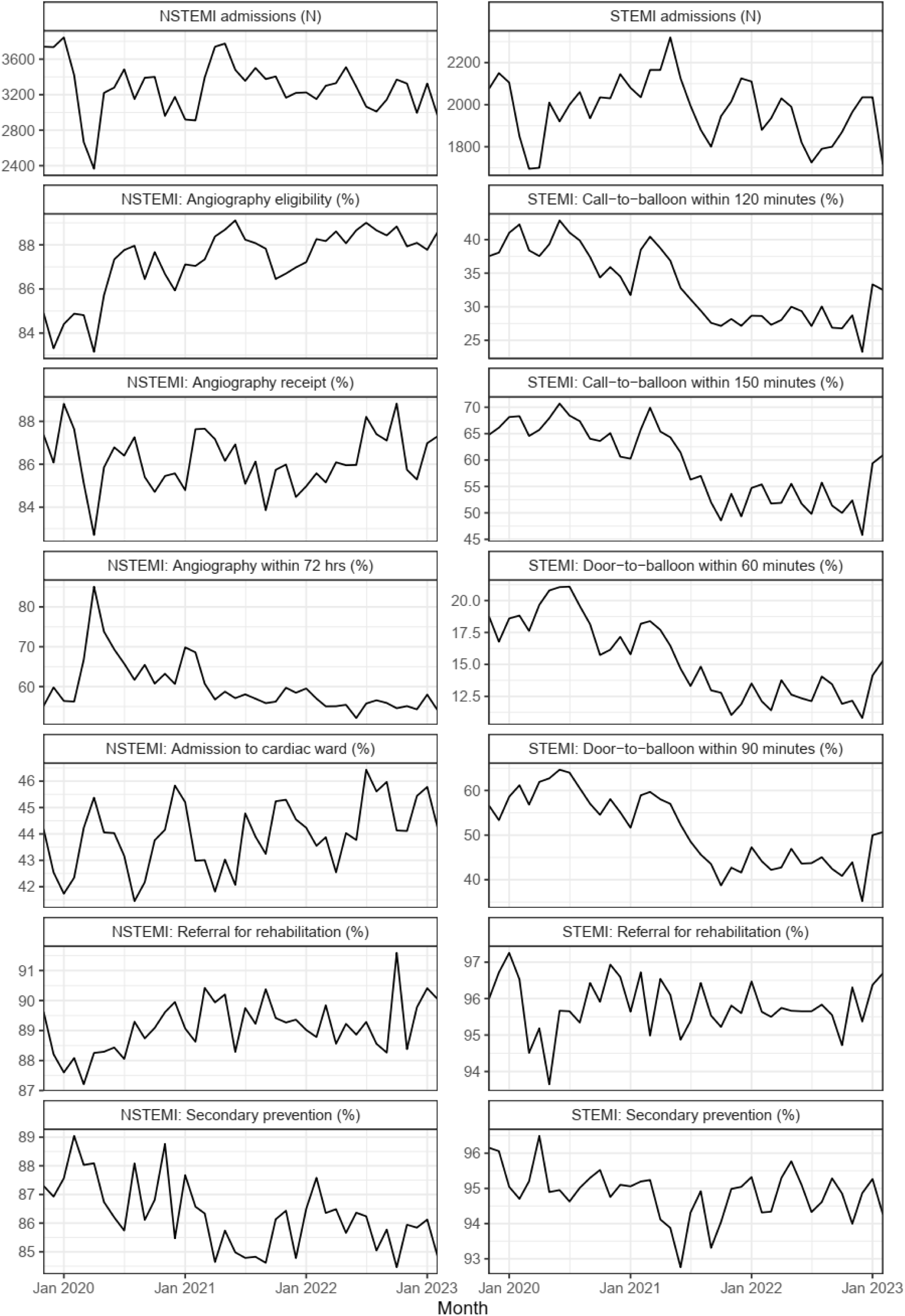
MI admissions and receipt of care standards by month

**Figure S7:**
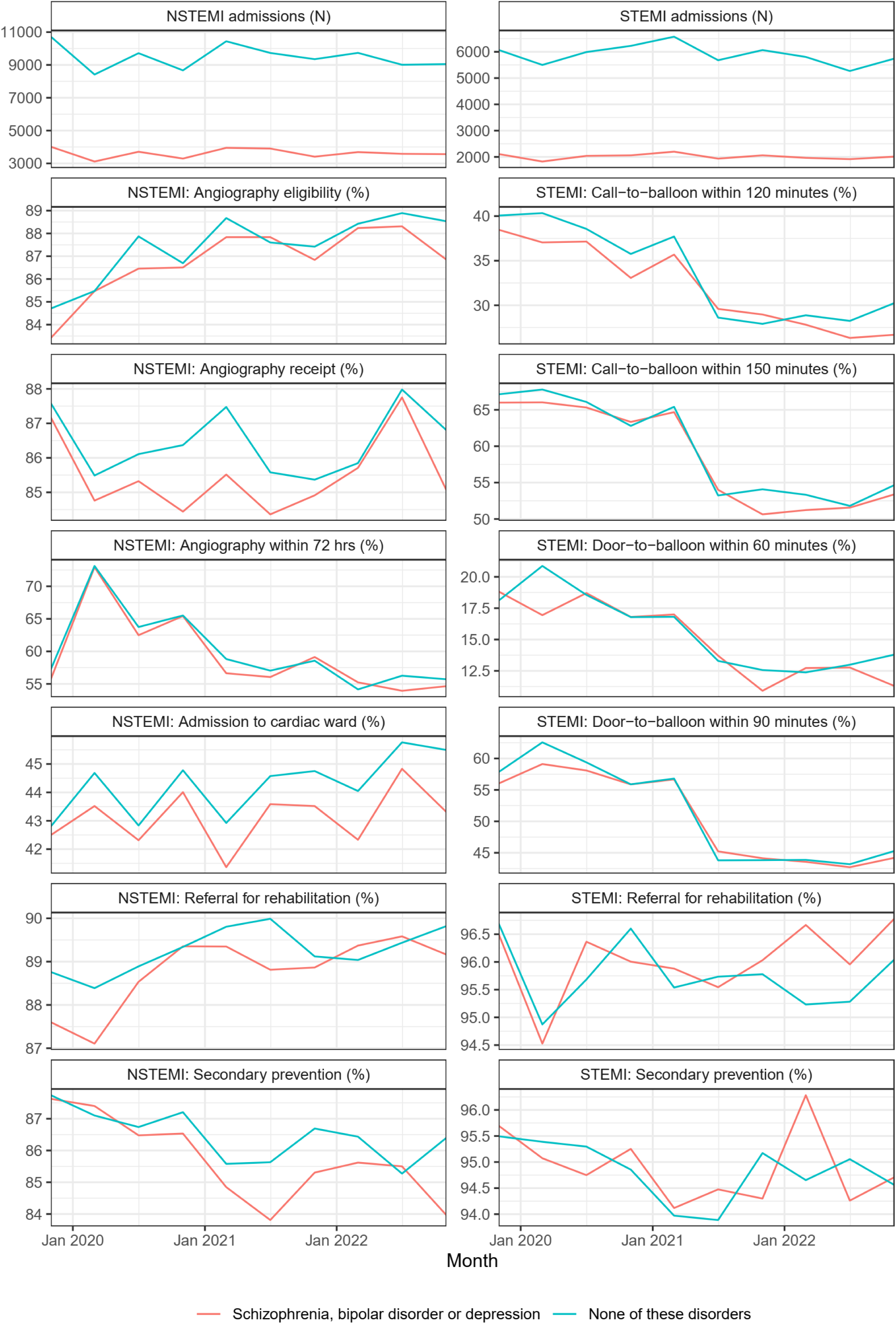
MI admissions and receipt of care standards by 4-month period and mental disorder

**Table S1:** Coronary intervention in patients who received angiography and have a MINAP record of an NSTEMI, by mental disorder status^a^.

| Characteristic | Schizophrenia<br>(N = 490) | Bipolar<br>disorder<br>(N = 510) | Depression<br>(N = 24 580) | None of these<br>disorders<br>(N = 68 575) |
| --- | --- | --- | --- | --- |
| **Coronary intervention amongst patients who received angiography** |  |  |  |  |
| PCI | 225 (45.9%) | 265 (52.0%) | 13 290 (54.1%) | 38 310 (55.9%) |
| PCI planned after discharge | <10 (<2.0%) | <10 (<2.0%) | 255 (1.0%) | 835 (1.2%) |
| CABG | 25 (5.1%) | 10 (2.0%) | 870 (3.5%) | 3220 (4.7%) |
| CABG planned after discharge | 15 (3.1%) | 15 (2.9%) | 650 (2.6%) | 2425 (3.5%) |
| Patient refused | <10 (<2.0%) | <10 (<2.0%) | 45 (0.2%) | 130 (0.2%) |
| Not performed or arranged | 95 (19.4%) | 80 (15.7%) | 3545 (14.4%) | 8685 (12.7%) |
| Not applicable | 45 (9.2%) | 50 (9.8%) | 2170 (8.8%) | 5635 (8.2%) |
| Missing | 70 (14.3%) | 75 (14.7%) | 3755 (15.3%) | 9340 (13.6%) |

CABG: coronary artery bypass graft, PCI: percutaneous coronary intervention.
<sup>a</sup>In accordance with NHSE's SDE statistical disclosure control rules, counts are rounded to the nearest 5 and counts less than 10 are suppressed

**Table S2:** Initial reperfusion treatment in patients with a MINAP record of a STEMI, by mental disorder status^a^.

| Characteristic | Schizophrenia<br>(N = 490) | Bipolar<br>disorder<br>(N = 405) | Depression<br>(N = 19 230) | None of these<br>disorders<br>(N = 58 920) |
| --- | --- | --- | --- | --- |
| **Initial reperfusion treatment** |  |  |  |  |
| pPCI in house | 340 (69.4%) | 310 (76.5%) | 15075 (78.4%) | 46400 (78.8%) |
| pPCI already performed at the interventional hospital | <10 (<2.0%) | <10 (<2.5%) | 260 (1.4%) | 680 (1.2%) |
| Referred for consideration for pPCI elsewhere | 15 (3.1%) | <10 (<2.5%) | 315 (1.6%) | 935 (1.6%) |
| Thrombolytic treatment | <10 (<2.0%) | <10 (<2.5%) | 60 (0.3%) | 125 (0.2%) |
| None | 120 (24.5%) | 85 (21.0%) | 3340 (17.4%) | 10210 (17.3%) |
| Missing | 10 (2.0%) | <10 (<2.5%) | 175 (0.9%) | 570 (1.0%) |
| **Reason treatment not given amongst patients where initial reperfusion treatment was recorded as 'None'** |  |  |  |  |
| N | 120 | 85 | 3340 | 10 210 |
| Ineligible ECG | 15 (12.5%) | <10 (<11.8%) | 350 (10.5%) | 1015 (9.9%) |
| Too late | 40 (33.3%) | 30 (35.3%) | 1020 (30.5%) | 3325 (32.6%) |
| Risk of haemorrhage | <10 (<8.3%) | <10 (<11.8%) | 25 (0.7%) | 50 (0.5%) |
| Uncontrolled hypertension | <10 (<8.3%) | <10 (<11.8%) | <10 (<0.3%) | <10 (<0.1%) |
| Administrative failure | <10 (<8.3%) | <10 (<11.8%) | 15 (0.4%) | 40 (0.4%) |
| Elective decision | 35 (29.2%) | 20 (23.5%) | 895 (26.8%) | 2485 (24.3%) |
| Patient refused treatment | <10 (<8.3%) | <10 (<11.8%) | 55 (1.6%) | 110 (1.1%) |
| None | <10 (<8.3%) | <10 (<11.8%) | 115 (3.4%) | 415 (4.1%) |
| Other | 20 (16.7%) | 15 (17.6%) | 690 (20.7%) | 2175 (21.3%) |
| Missing | <10 (<8.3%) | <10 (<11.8%) | 165 (4.9%) | 590 (5.8%) |

pPCI: primary percutaneous coronary intervention
<sup>a</sup>In accordance with NHSE's SDE statistical disclosure control rules, counts are rounded to the nearest 5 and counts less than 10 are suppressed

**Table S3:**
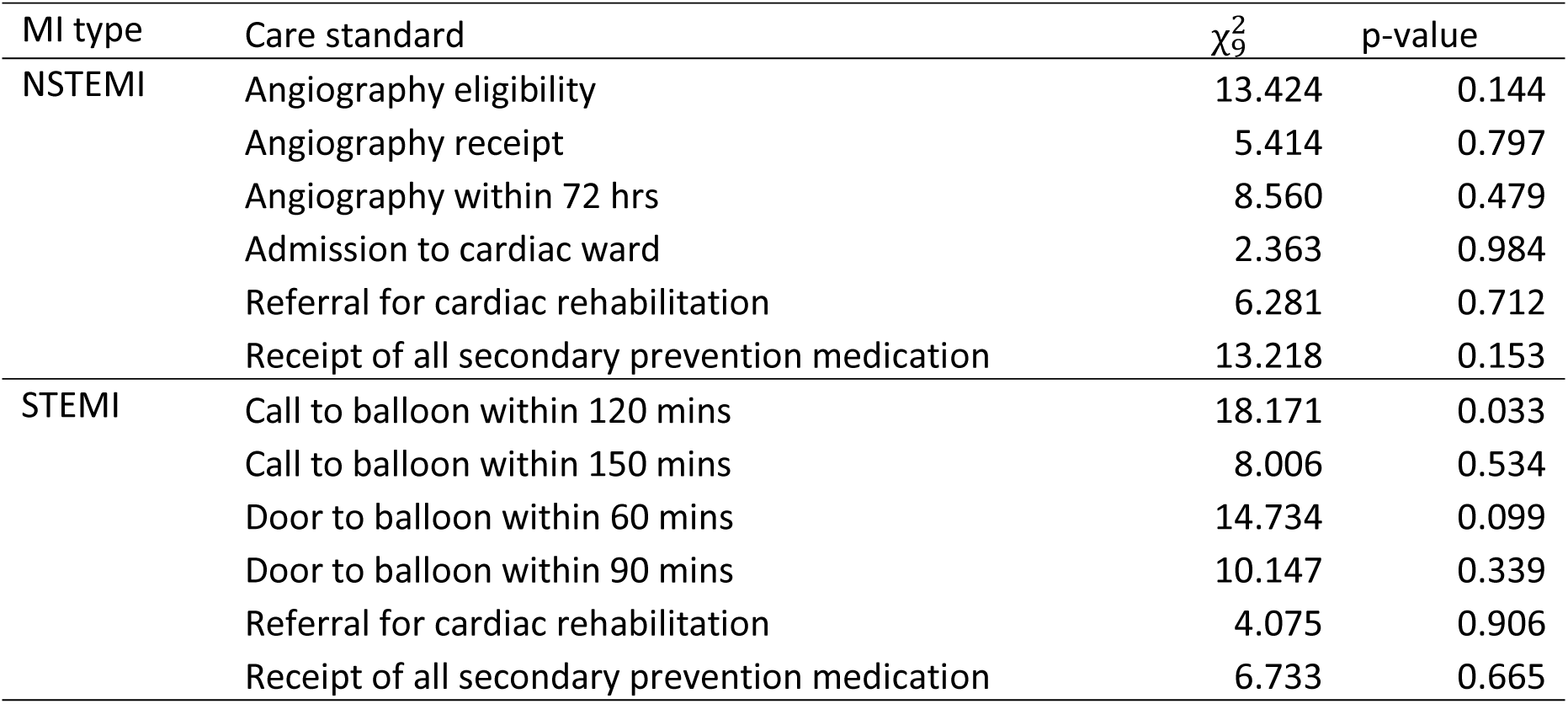
Interaction between mental disorder and 4-month time period; likelihood ratio tests of whether adding the interaction improved the fit of the model (primary analysis)

| MI type | Care standard | $\chi^2_9$ | p-value |
| --- | --- | --- | --- |
| NSTEMI | Angiography eligibility | 13.424 | 0.144 |
|  | Angiography receipt | 5.414 | 0.797 |
|  | Angiography within 72 hrs | 8.560 | 0.479 |
|  | Admission to cardiac ward | 2.363 | 0.984 |
|  | Referral for cardiac rehabilitation | 6.281 | 0.712 |
|  | Receipt of all secondary prevention medication | 13.218 | 0.153 |
| STEMI | Call to balloon within 120 mins | 18.171 | 0.033 |
|  | Call to balloon within 150 mins | 8.006 | 0.534 |
|  | Door to balloon within 60 mins | 14.734 | 0.099 |
|  | Door to balloon within 90 mins | 10.147 | 0.339 |
|  | Referral for cardiac rehabilitation | 4.075 | 0.906 |
|  | Receipt of all secondary prevention medication | 6.733 | 0.665 |

**Table S4:** Odds ratios by 4-month time period for call-to-balloon within 120 mins following STEMI in patients with versus without any of schizophrenia, bipolar disorder or depression. Model 5 with interaction between mental disorder and 4-month time period.

| 4-month time period | Odds ratio (95% confidence interval) |
| --- | --- |
| Nov 2019 - Feb 2020 | 0.92 (0.80, 1.06) |
| Mar 2020 - Jun 2020 | 0.97 (0.84, 1.13) |
| Jul 2020 - Oct 2020 | 1.01 (0.87, 1.16) |
| Nov 2020 - Feb 2021 | 0.85 (0.73, 0.98) |
| Mar 2021 - Jun 2021 | 0.93 (0.81, 1.06) |
| Jul 2021 - Oct 2021 | 1.15 (0.99, 1.35) |
| Nov 2021 - Feb 2022 | 1.11 (0.95, 1.29) |
| Mar 2022 - Jun 2022 | 1.00 (0.85, 1.16) |
| Jul 2022 - Oct 2022 | 1.03 (0.88, 1.21) |
| Nov 2022 - Feb 2023 | 0.81 (0.68, 0.95) |

**Table S5:** Interaction between mental disorder and 4-month time period; likelihood ratio tests of whether adding the interaction improved the fit of the sensitivity analysis models.

| MI type | Care standard | $\chi^2_9$ | p-value |
| --- | --- | --- | --- |
| NSTEMI | Angiography eligibility | 13.076 | 0.159 |
|  | Angiography receipt | 6.143 | 0.726 |
|  | Angiography within 72 hrs | 6.234 | 0.716 |
|  | Admission to cardiac ward | 2.421 | 0.983 |
|  | Referral for cardiac rehabilitation | 3.408 | 0.946 |
|  | Receipt of all secondary prevention medication | 10.922 | 0.281 |
| STEMI | Call to balloon within 120 mins | 9.678 | 0.377 |
|  | Call to balloon within 150 mins | 6.409 | 0.698 |
|  | Door to balloon within 60 mins | 14.482 | 0.106 |
|  | Door to balloon within 90 mins | 10.608 | 0.304 |
|  | Referral for cardiac rehabilitation | 4.465 | 0.878 |
|  | Receipt of all secondary prevention medication | 10.861 | 0.285 |

## The RECORD statement – checklist of items, extended from the STROBE statement, that should be reported in observational studies using routinely collected health data.

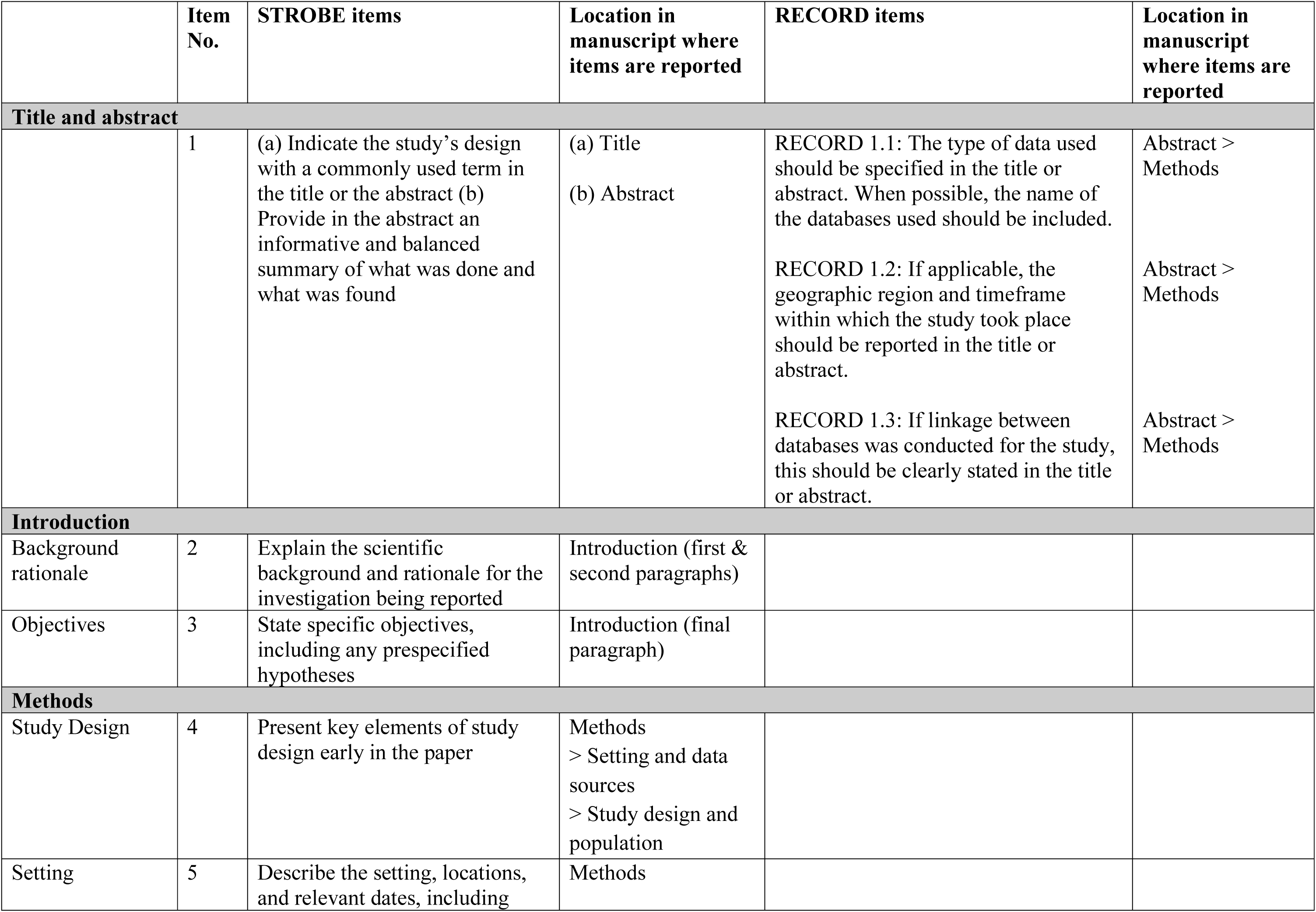
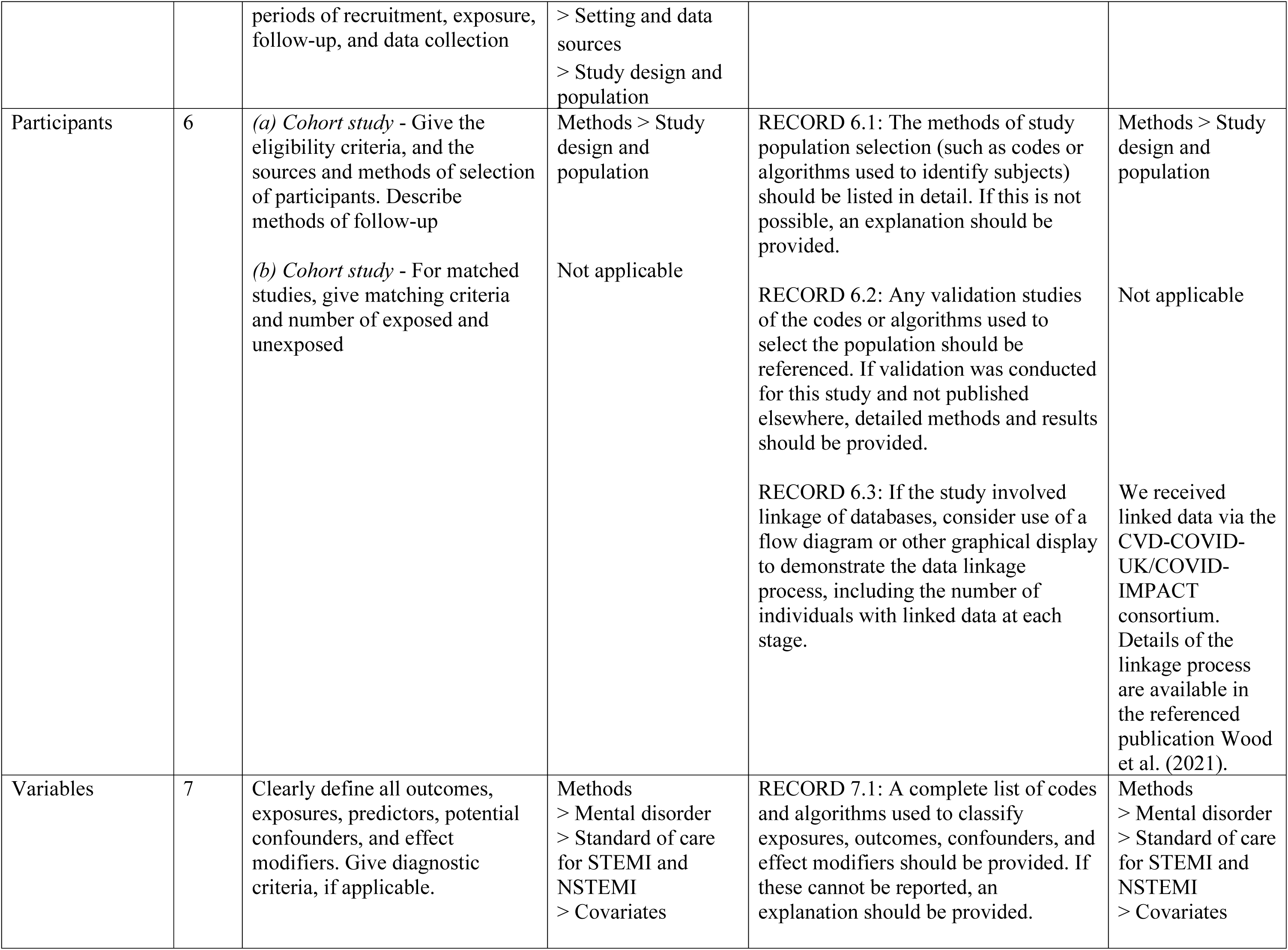
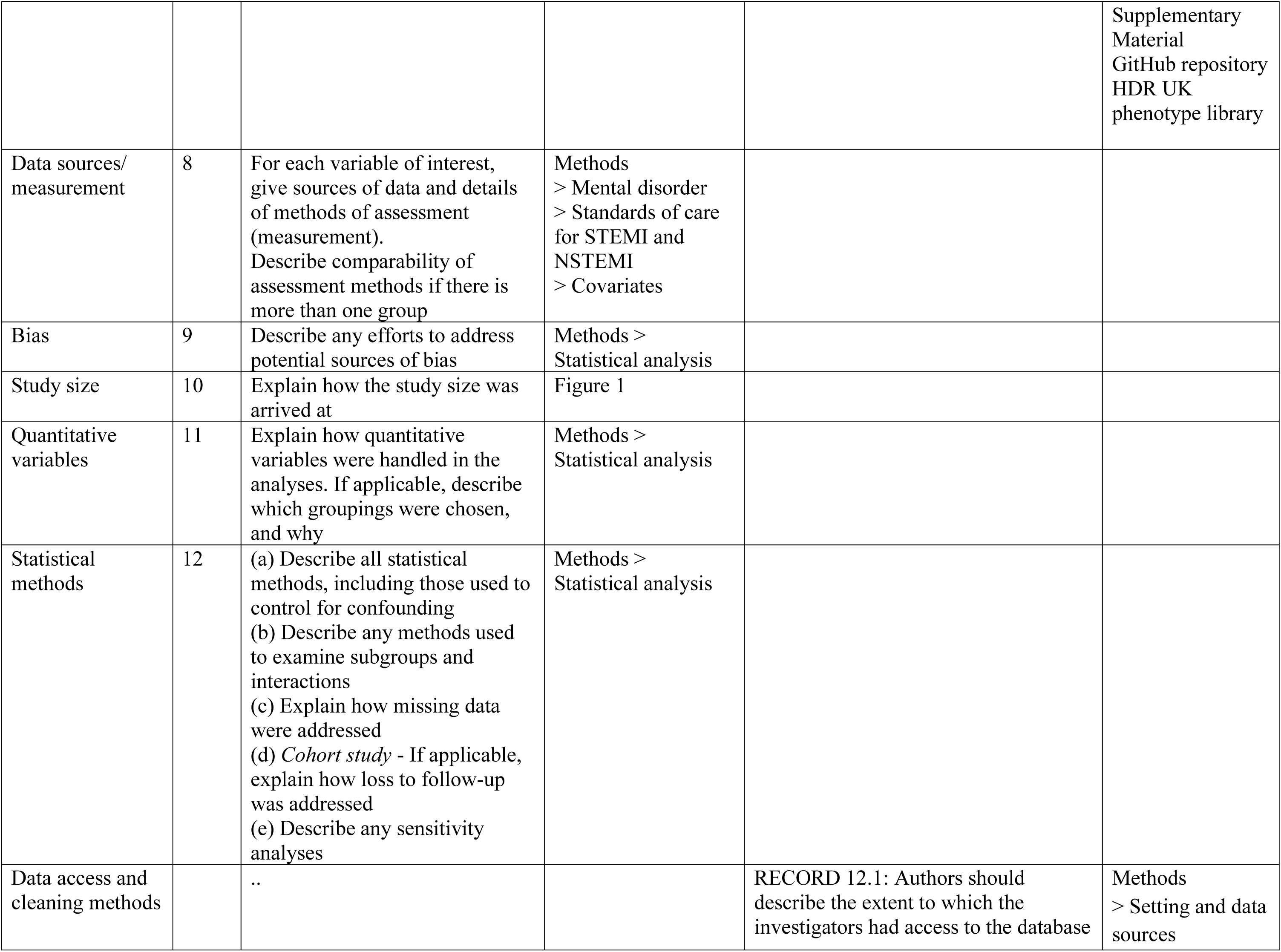
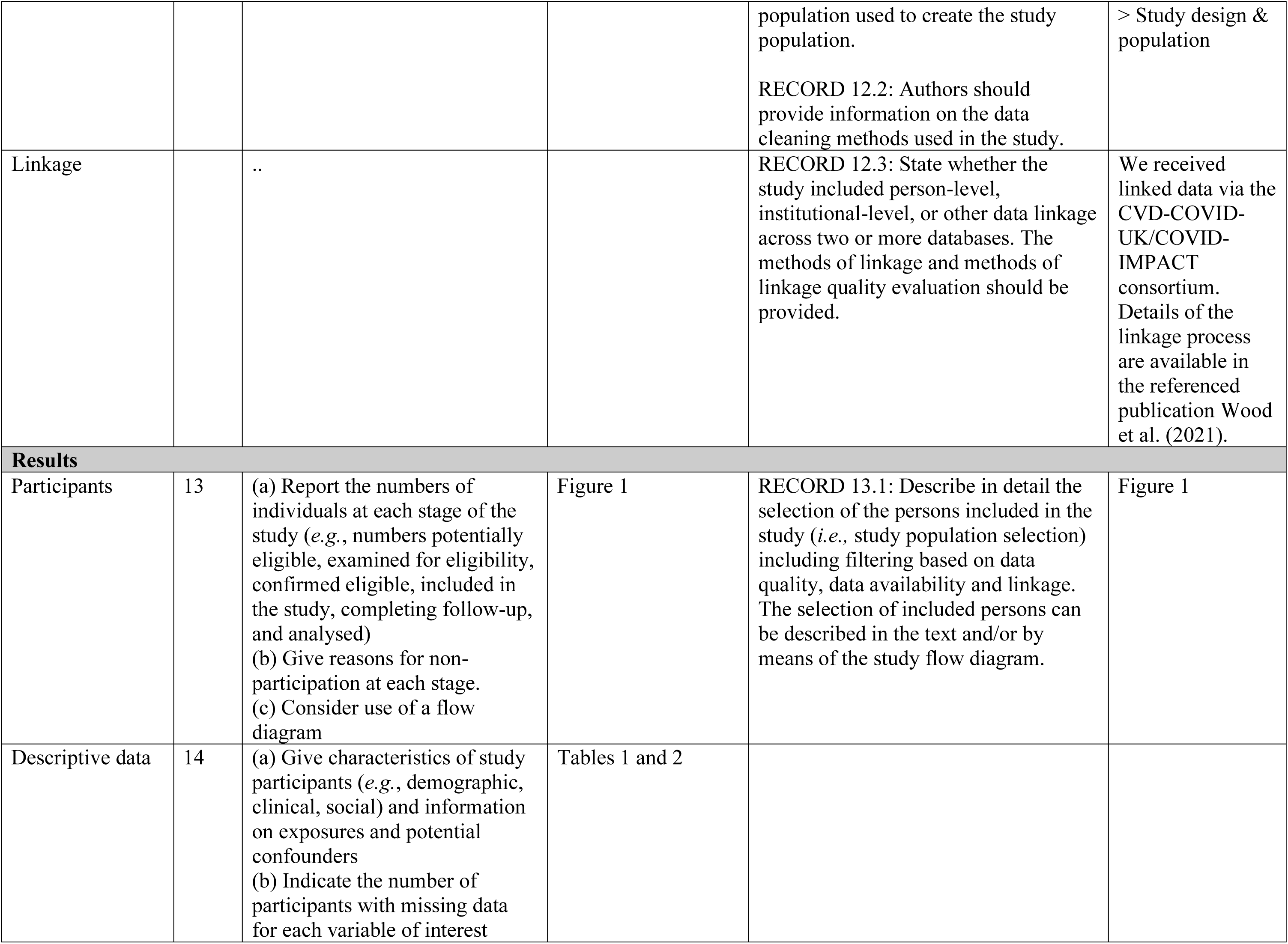
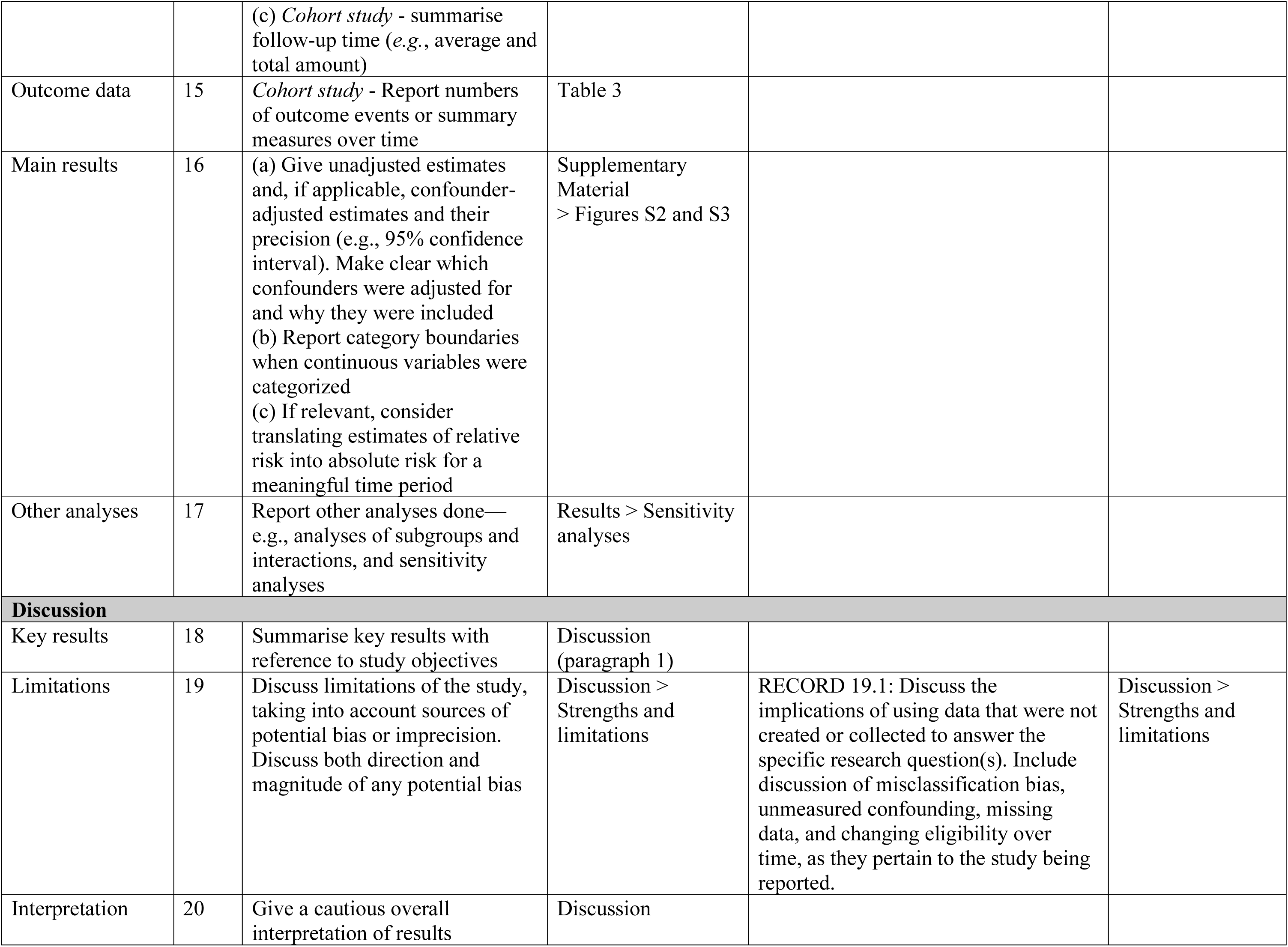
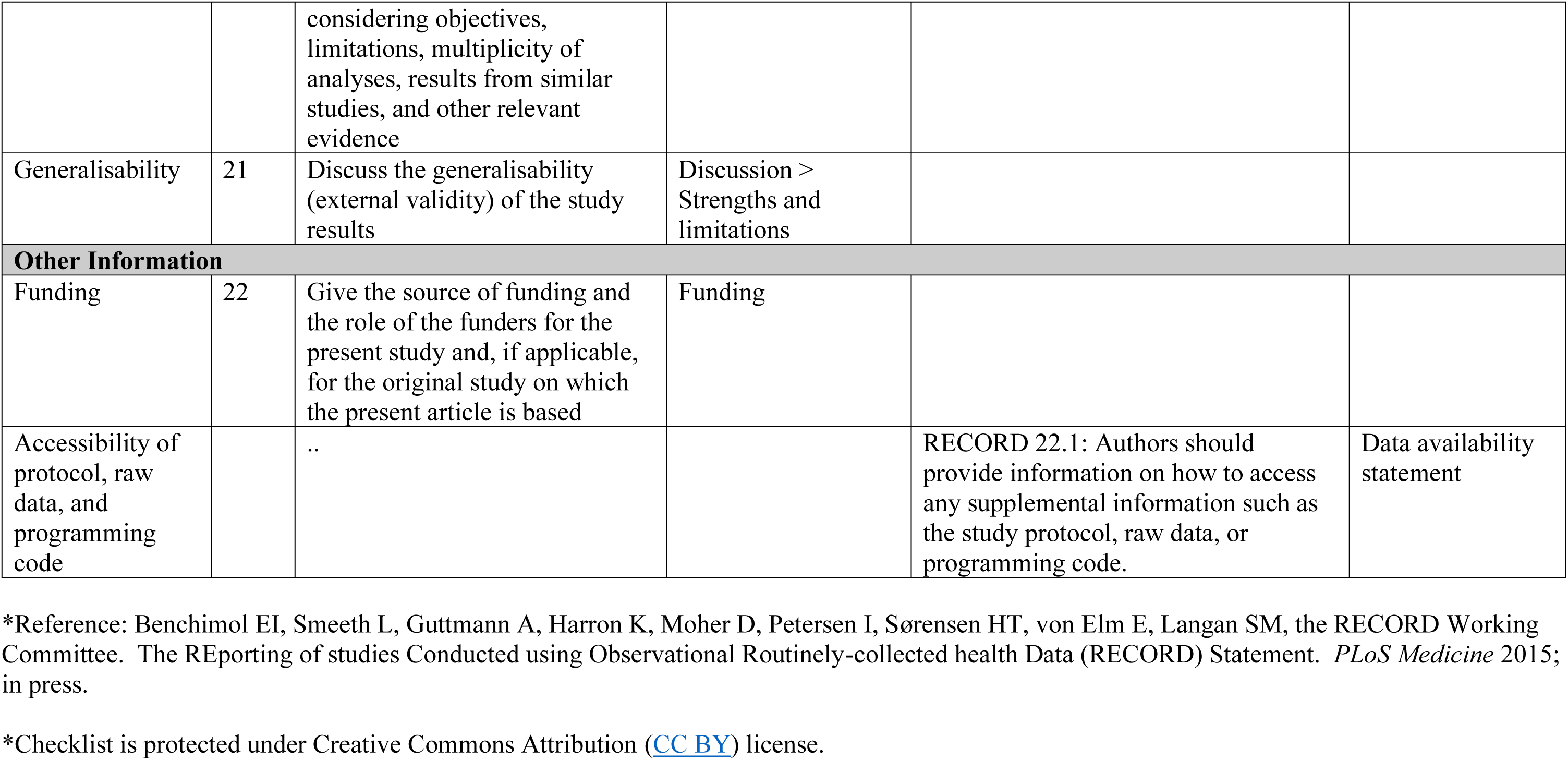

## Notes

### Clinical Protocols

https://github.com/BHFDSC/CCU046_01

### Author Declarations

The North East - Newcastle and North Tyneside 2 research ethics committee provided ethical approval for the CVD-COVID-UK/COVID-IMPACT research programme (REC No 20/NE/0161) to access, within secure trusted research environments, unconsented, whole-population, de-identified data from electronic health records collected as part of the routine healthcare of patients.

